# Returning Research Results That Indicate Risk of Alzheimer Disease Dementia to Healthy Participants in Longitudinal Studies (WeSHARE)

**DOI:** 10.1101/2024.07.01.24309801

**Authors:** Sarah M. Hartz, Spondita Goswami, Amy Oliver, Alissa Evans, Sacha Jackson, Erin Linnenbringer, Krista Moulder, John C. Morris, Jessica Mozersky

**Affiliations:** Washington University in St. Louis School of Medicine; Washington University in St Louis

## Abstract

**Introduction:** Returning research results that indicate risk of Alzheimer disease (AD) - a disease for which no meaningful treatments or cure exist - to cognitively normal participants is controversial. AD is thought to begin many years before clinical signs and symptoms begin. During this time, individuals are cognitively normal, but have biomarkers that indicate pathophysiological changes in the brain. With this study, we aim to evaluate impact of returning research results on cognitively normal participants recruited from a longitudinal observational cohort on aging at the Knight Alzheimer Disease Research Center (Knight ADRC) at Washington University in St. Louis.

**Methods and analysis:** Our study uses a 2-year, delayed-start randomized clinical trial design. Participants are randomized to receive their research results either in 2-4 weeks or 1 year after informed consent. We plan to approach approximately 260 participants who have research results from previous genetic and biomarker testing at the Knight ADRC. The primary cognitive outcomes are 1-year change in subjective cognitive score (Clinical Dementia Rating (CDR^®^) sum of boxes), objective cognitive score (psychometric composite score), and the primary psychosocial outcome is Impact of Event-Revised score 1 year after return of research results.

**Ethics and dissemination:** This study has been approved by the WUSM Institutional Review Board (IRB) and the Human Research Protection Office (HRPO). Results from these trials are shared through conferences and publications.

**Trial registration number:** NCT04699786.

**Strengths and limitations of this study:**

- This study empirically evaluates how returning research AD biomarkers impacts participants both psychologically and cognitively using a randomized delayed-start clinical trial
- Multiple research AD biomarkers are synthesized into a single estimate for AD dementia risk
- A delayed-start design is necessary because it is unethical to withhold research results from participants but may limit the generalizability of the results because all participants will be receiving their research results

## INTRODUCTION

Alzheimer disease is a progressive neurodegenerative disorder characterized by a prolonged preclinical phase before significant cognitive or behavioral symptoms emerge. The clinical manifestation of the disorder is characterized by cognitive deterioration, changes in behavior and increased requirement for care due to loss of functional independence. Before these signs appear, the brain undergoes structural and functional changes triggered by biomarkers such as amyloid beta (Aβ) accumulation and neurofibrillary tangle pathology. AD biomarkers can be detected years before any clinical symptoms emerge, known as the preclinical phase.^1–3^ This preclinical phase, characterized by biomarker evidence of AD neuropathology, provides an opportunity for potential clinical intervention, thereby improving our understanding of the disease.

There is considerable interest in learning AD biomarker status among research participants,^4–8^ although limited data exist related to the impact of disclosing such research results to cognitively normal participants, especially those enrolled in longitudinal studies where there may not be any immediate actionability of knowing one’s biomarker status such as entering a clinical trial. Current research efforts are focused on identifying and characterizing biomarkers that are associated with the onset of AD dementia or an increased risk of developing AD dementia, and often utilize longitudinal cohorts that enable individuals to be followed over time. Such research specifically aims to enroll individuals with normal cognitive function but who exhibit brain pathology or biomarkers that indicate brain pathophysiological changes that increase risk of developing AD.^3, 9–14^

Ethical considerations arise when disclosing biomarker research results to cognitively normal participants due to concerns about the potential negative consequences arising from disclosure. These concerns include psychological harms such as anxiety or depression and spurious cognitive test scores due to subjective feelings of improved or worsening memory arising from the knowledge of personal AD risks. However, evidence from studies to date does not suggest major psychological harms following disclosure although long-term follow up is limited.^15–22^ Conversely, there may be benefits to disclosure such as increased retention, particularly among underrepresented populations, and increased satisfaction with study visits^23–26^.

In addition, most studies examine psychological outcomes following disclosure of a single result – such as amyloid PET status – and there is a lack of data on the impact of learning about multiple types of risk factors such as genetic and neuroimaging results^7^. There is also no data on uptake and who declines to learn their AD research results.

Given the potential benefits of sharing research results with participants and the high stated interest among participants, it is crucial to investigate the impact of disclosure on outcomes and understand who opts in and out of return of research results (RoRR). This study aims to bridge this gap by building a protocol that (1) discloses research results that include multiple biomarkers and a synthesis of absolute risk^27^ to participants while measuring the psychological and cognitive effects on cognitively normal individuals enrolled in a longitudinal cohort of aging, and (2) delineates steps to examine the actual uptake of research results, including who declines to learn their research results and the process of qualitatively evaluating the reasons behind participants’ decisions to receive/decline their results. This study was designed to create an evidence base regarding the optimal way of disclosing estimated risk based on combined biomarker results, essential as clinical breakthroughs such as disease modifying therapies emerge and therefore necessitate disclosure of biomarker status. Here, we describe a protocol for returning AD biomarker research results and evaluating the impact of disclosure on psychological, behavioral, and cognitive outcomes.

These findings will help provide a framework for safely, ethically, and respectfully disclosing research results that honor participants’ autonomy.

## METHODS AND ANALYSIS

The trial’s objective is to identify the most effective approach for RoRR to participants, incorporating personalized 5-year AD dementia risk combining predictions based on *APOE* genotyping, recent imaging (amyloid PET and MRI), age, gender, and race. The aim is to develop a process that minimizes potential psychological and behavioral outcomes and cognitive decline resulting from learning these results. We will use a delayed-start randomized noninferiority clinical trial to determine the impact of returning research results that indicate AD dementia risk on cognitive and psychosocial outcomes (**Figure 1**).

**Figure 1:**
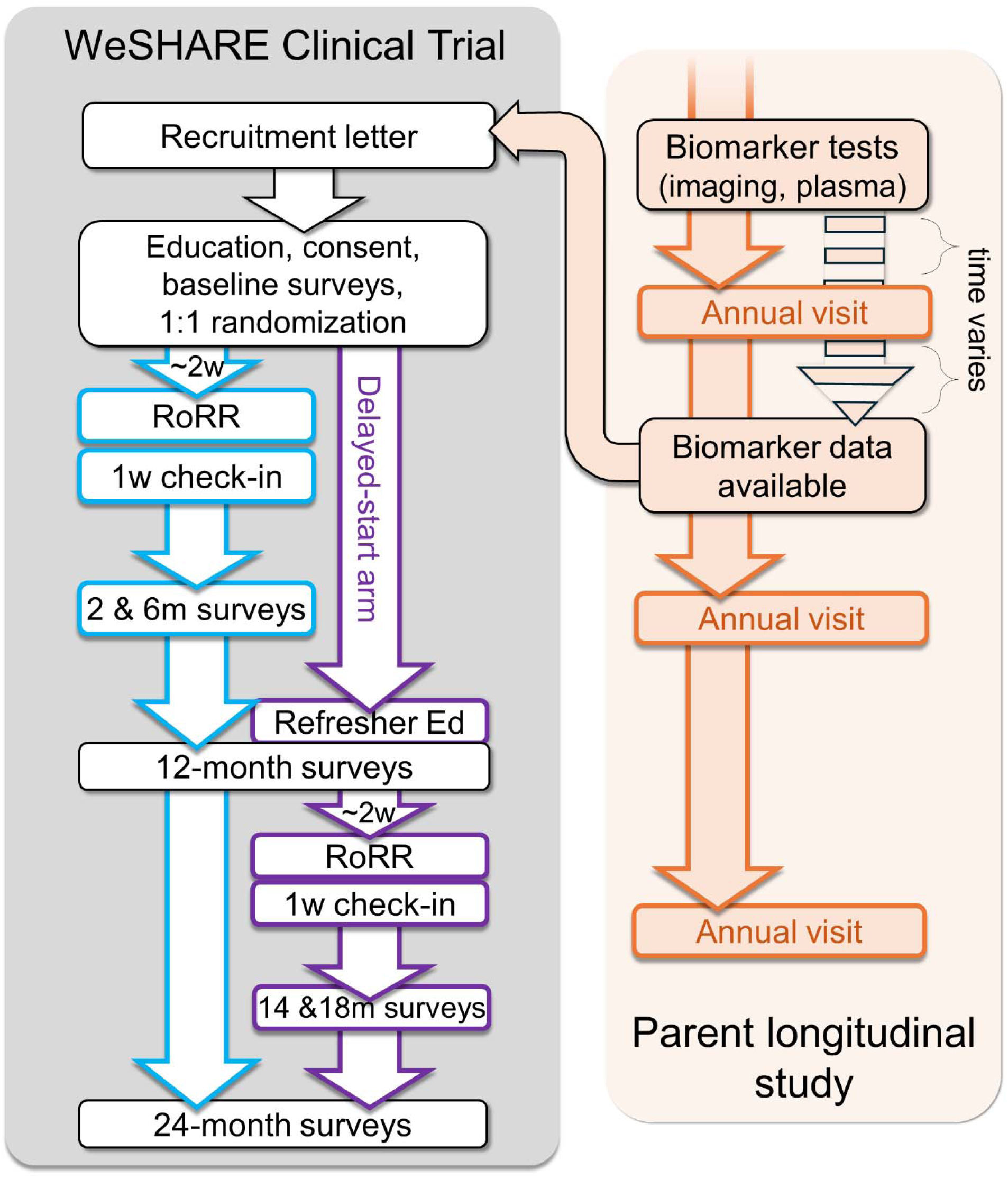
Design of WeSHARE clinical trial and its relationship to the parent longitudinal, observational study (Memory and Aging Project at the Knight ADRC).

Given the limited literature on informing research participants about their estimated AD symptom risk, the hypothesis is that any significant cognitive or psychosocial changes will normalize within a year. Consequently, individuals in the RoRR arm are not expected to differ substantially from those in the delayed-start arm at the 12-month mark, just before the delayed-start arm receives their results.

### Inclusion and exclusion criteria

Participants eligible for inclusion in the study will need to meet the following criteria: (1) current Knight ADRC participant that consented to be contacted for other research opportunities, (2) classified as cognitively normal (Clinical Dementia Rating^28, 29^ (CDR®) = 0) within the past 12 months, (3) aged 65 years or older, (4) available research *APOE* genotype, and (5) either recent research brain MRI and amyloid PET scan results, or recent plasma amyloid measurements (ideally within the past two years, but up to five years will be acceptable due to COVID-19-related delays). There are no specific exclusion criteria.

### Participant contact

Prior to scheduling the annual cognitive visit, eligible participants will be mailed a letter informing them of their eligibility, educational materials (Appendix), and a consent form to review. Approximately two weeks after the study packets are mailed, a study team member will call each potential participant to ask if they have any additional questions based on the information they received and whether they are interested in participating. If participants agree, they will be scheduled for their first visit. A study coordinator will conduct the first visit over Zoom-HIPAA or in person at the Knight ADRC. During this visit, the study coordinator will go through the educational materials, examples of research results, and their limitations, and will obtain informed consent. The educational materials will provide information on the risk factors associated with AD dementia, including age, family history, gender, and educational attainment. Additionally, other risk factors, such as those identified through research, will be introduced, with a reminder that having these factors does not guarantee developing AD dementia. Participants will be informed about the possibility of receiving increased or not increased risk, including explanation of what these results signify and discussion of their limitations. Emphasis will be placed on the fact that the research risk estimate predicts the likelihood of experiencing early symptoms of AD dementia.

The informed consent process will be completed if the participant chooses to move forward. After enrollment, using a delayed-start randomized clinical trial design, participants will be randomized to intervention or delayed-start (receiving results in approximately 1 year). After consent from participants during Visit 1, the intervention arm will be scheduled immediately for RoRR in Visit 2 with a clinician for disclosure, whereas the delayed start arm will schedule Visit 2 one year later.

### Disclosure process

At Visit 2 a clinician will show the participant their research results report (example in Appendix), explain the report and answer any questions (training manual for RoRR in Appendix). Specifically, participants learn about three key pieces of information. First, their baseline absolute risk of developing AD dementia will be addressed, considering their demographic characteristics. Then, the disclosure process will shift to the adjusted absolute risk of AD dementia based on the research results, with a direct comparison made between the adjusted result and the baseline result. This will include an explanation that the range of adjusted risk represents the best estimates derived from current data. Following this, a review of the individual research results contributing to the adjusted risk will be discussed. This will include discussing biomarker test results (such as amyloid PET and MRI or an amyloid blood test) and the *APOE* genetic test result. Finally, there will be room for additional discussion and questions to ensure the participant comprehensively understands and can engage with the information provided. Disclosure sessions will be scheduled for approximately 30 minutes to allow time for questions and education, and the duration will be tracked. Participants will receive a call from the research coordinator one week after receiving results to answer any questions or concerns that may have come up.

### Data collection

Data will be collected through surveys using validated measures and semi-structured interviews with participants. Specifically, semi-structured interviews will be conducted with participants who either received or opted out of receiving research results. Surveys will be employed for quantitative assessments of subjective measures of cognitive and psychological outcomes and objective measure of cognitive outcome at baseline, 2 months, 6 months, 12 months, and 24 months. Participant counts will be utilized to determine the number of participants who opted to either receive or not receive research results.

Survey instruments include published quantitative measures (including decision to receive results, objective and subjective cognitive scores, impact of event, AD related distress, decisional regret, depression, feelings about results, healthcare utilization, lifestyle/health behavior changes, satisfaction with RoRR, personal utility, and comprehension of results). The schedule for survey administration is given in Table 1. To ensure a satisfactory response rate, surveys will be offered via multiple modalities, including phone, email and postal mail.

**Table 1:**
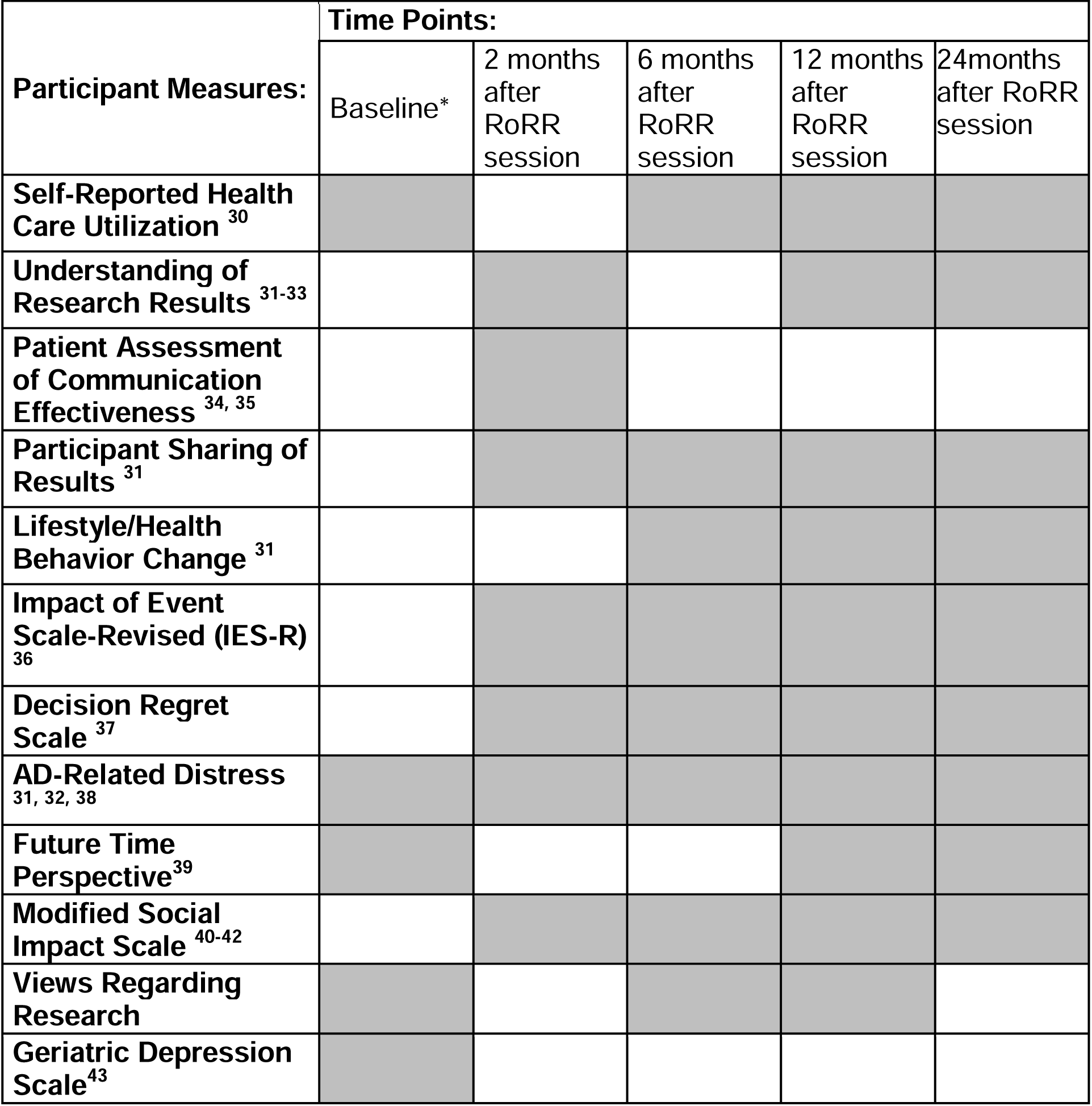
Schedule of outcomes and measures in quantitative surveys and constructs in qualitative Interviews.

#### Data analysis

We hypothesize that significant cognitive or psychosocial changes will normalize after one year, leading to no substantive clinical differences between individuals in the RoRR arm and those in the delayed-start arm at the 12-month time point (prior to the delayed-start arm receiving results). We will analyze cognitive or psychosocial changes based on RoRR, stratifying by whether the participant received increased risk results. The primary cognitive outcomes include the 1-year change in subjective cognitive score (CDR® sum of boxes) and objective cognitive score (psychometric composite), while the primary psychosocial outcome is the impact of event score at 1 year. Our primary analyses will involve logistic regressions comparing the RoRR arm to the delayed-start arm at 12 months, stratified by risk (increased risk vs. no change in risk or decreased risk) and adjusted for age, gender, and race. Secondary statistical analyses will use logistic and/or linear regressions to determine the effect of RoRR on the other outcomes (see Table 2). We will also explore changes in participation rates potentially due to RoRR and the impact of RoRR on available study partners/families in terms of decisional regret and feelings about the result.

### Psychological support

Given concerns about the potential for adverse psychosocial outcomes of RoRR, study staff interacting with participants will notify a board-certified psychiatrist (one of the two principal investigators of this study) with any concerns. Any unanticipated adverse events will be reported to the IRB, and all adverse events (anticipated or unanticipated, serious or not, related or unrelated) will be reported to the funding agency. Based on prior studies of return of results, we do not anticipate any new-onset depression or suicidality, although it is important to ensure that this is monitored.

## Discussion

Returning AD dementia risk results to cognitively normal participants has become increasingly important due to several factors, such as maintaining participant autonomy, improving clinical care, and including high-risk participants in clinical treatment studies. Currently, there is limited evidence regarding psychological and cognitive impact of RoRR on participants. Therefore, data is needed to inform this discussion and shape policies, protocols, and clinical care. This longitudinal delayed-start noninferiority study aims to address the evidentiary gap. Psychological and cognitive data will allow for comparisons between individuals who receive results (either immediately or on a delay) and those who decline to receive results.

This study is the first to return multiple types of research results - genetic and biomarker-based - that indicate the risk for AD dementia. Prior studies have evaluated the impact of returning a single research result in specific contexts, but no studies have evaluated the impact of returning multiple research results synthesized into a single estimate for AD dementia risk, which will be necessary as precision medicine evolves and data volume increases. Additionally, this study will provide evidence on the actual uptake of AD dementia risk research results in a real-world setting, considering the potential gap between intentions and behaviors observed in prior genetic testing contexts.. Understanding the impact of RoRR on cognitive outcomes will inform future study designs. Lastly, we will develop educational materials and a training module for healthcare providers based on existing protocols used in AD settings, adapting and expanding these protocols to include different types of research results.

We plan to return research, not clinical, results that meet the quality threshold recommended by the National Academy of Sciences (NAS). NAS guidelines state that laboratory analysis must provide confidence in the result, especially when results are not intended for clinical decision-making, as in our study. Our research uses state-of-the-art imaging and genotyping technology to ensure quality and accuracy. Results will be presented as a single risk estimate representing a 5-year risk of developing symptomatic AD dementia, incorporating published AD incidence rates and risk curves. This flexible approach can accommodate new biomarkers as evidence emerges.

### Dissemination

We plan to continue to share the results of this trial through local, national and international conferences and publications.

## Data Availability

All data produced in the present study are available upon reasonable request to the authors

