## Supplemental material for "Returning Research Results That Indicate Risk of Alzheimer Disease Dementia to Healthy Participants in Longitudinal Studies (WeSHARE)"

Educational brochure (Imaging)

KnightADRC Alzheimer's Disease Research Center | WASHINGTON UNIVERSITY ST. LOUIS

For participants of the Knight Alzheimer Disease Research Center Memory and Aging Project, including the Adult Children Study

### Should I find out my research results related to Alzheimer disease dementia?

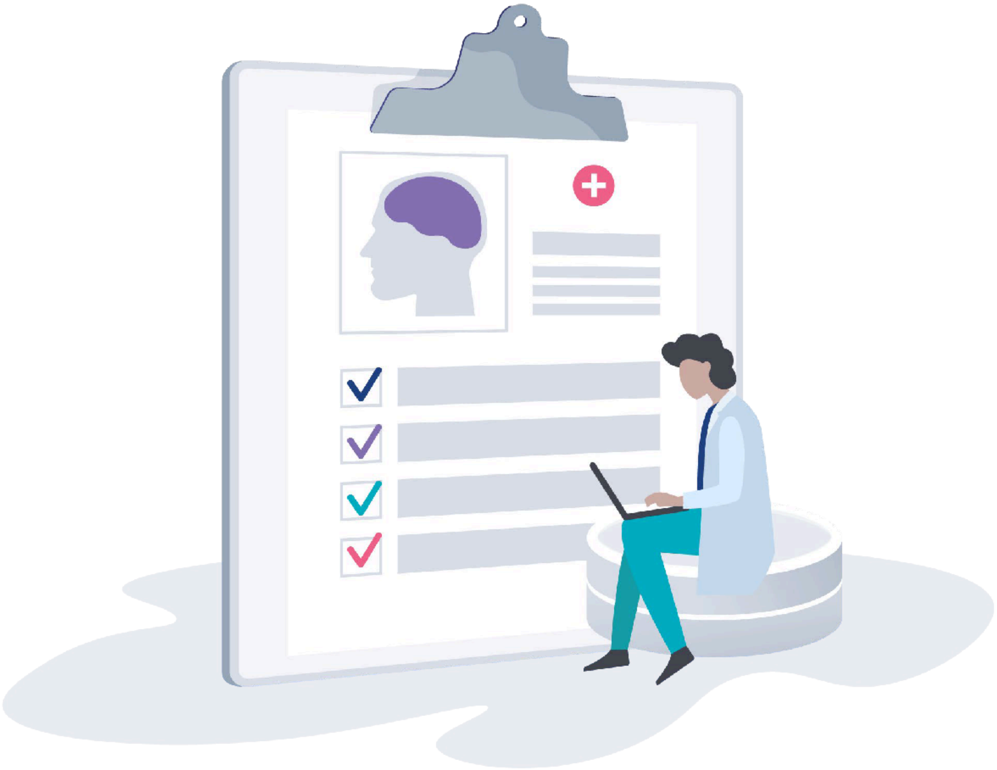

#### This guide will help you:

**Learn** about Alzheimer disease (AD) biomarkers, risk factors, and

**Know** what doctors and researchers know about risk of AD dementia

**Decide** if you want to see research results showing your risk of getting AD dementia

**Call** us if you want to see your research results or want more information

### How to use this guide

This guide can help you decide if you want to learn some of your brain imaging and blood test results. These results are from tests you had for the Memory and Aging Project (MAP) at the Knight Alzheimer Disease Research Center (ADRC).

Your results allow us to estimate your chance of developing early symptoms of Alzheimer disease (AD) dementia (problems with memory and thinking) in the next 5 years.

Your research results only give you an estimate of your risk. They show if you have factors that make developing AD dementia more, or less, likely. There is no information that can definitely tell if you will or won't get AD dementia.

Thank you for taking part in this AD research. Your help is important.

---

Please read this guide and use the decision worksheet on pages 6-7. This will help you decide if you would, or would not, like to see an estimate of your risk of developing AD dementia based on some of your research results.

If you are interested in getting your results, or would like more information, call the study team at 314-362-6535.

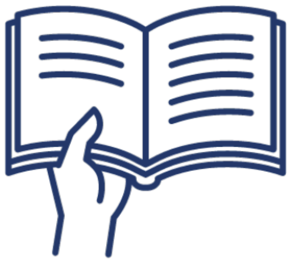

#### What's in this guide?

### What is Alzheimer disease (AD) dementia?

Dementia is the loss of memory and thinking abilities that interferes with a person's usual activities. These activities include things like driving, cooking or managing finances. There are many different types of dementia, but Alzheimer disease (AD) dementia is the most common type of dementia. Once the memory and thinking problems of AD dementia begin, they get worse over time. This makes it harder for a person to carry out daily activities. There is currently no cure or prevention for AD dementia. There are only treatments to help some of the symptoms.

### What are the risk factors for AD dementia?

AD risk factors are certain things about a person that can raise their chance of getting AD dementia. AD biomarkers and genes are risk factors for AD dementia. Having AD dementia risk factors doesn't necessarily mean you'll develop AD dementia. But, your chances are higher if you have certain risk factors.

#### AD biomarkers

Every person with AD dementia has a buildup of 2 proteins in their brain, called amyloid and tau. These proteins build up in the brain over many years before they cause any memory and thinking problems. Not everyone who has this buildup will develop AD dementia. AD researchers want to better understand why some people with buildup of these proteins develop AD dementia, while others don't.

To study AD dementia, researchers at the Knight ADRC use brain scans (PET scans and MRI), spinal fluid, and blood tests to measure these proteins and how they affect the brain. Measurements of amyloid, tau, or other AD dementia-related brain changes are AD biomarkers. Researchers have learned that:

- Not all people who show these AD biomarkers will develop AD dementia. Some people with AD biomarkers will never get memory and thinking problems.
- Some people who do not have any of these AD biomarkers now may still develop memory and thinking problems later.

#### The *APOE* gene

The *APOE (apolipoprotein E)* gene has three forms: *ε2, ε3,* and *ε4*. We all have two copies of the *APOE* gene. Each parent gives us one copy. Only *APOE ε4* makes it more likely that a person will get AD dementia. A blood test can tell which forms of *APOE* you have.

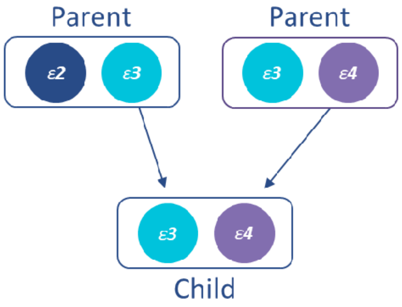

### What do researchers know about other risk factors for Alzheimer disease (AD) dementia?

**Having AD dementia risk factors does not always mean that you will develop AD dementia.** It is possible that a person with many AD risk factors will never develop AD dementia. Another person without AD risk factors might still develop AD dementia in the future.

#### These are the factors that increase anyone's risk of developing AD dementia:

**Age:** **The greatest risk factor for AD dementia is being older.** The older you are, the higher the chance you may develop AD dementia. About 1 in 3 (33%) older adults has AD dementia by the age of 85.

**Gender:** About 2 in 3 (or 66%) Americans with AD dementia are women. This difference is partly because women live longer than men. Older age is the greatest risk factor for developing AD dementia.

**Family history:** Your risk of AD dementia is higher if family members like a parent, sister, or brother have had it.

Even if your parent had AD dementia, this does not mean that you will develop AD dementia.

**Social Determinants of Health:** In the United States, access to healthcare, housing, food, education, and employment impacts people's health. Not having equal access to these necessities increases the risk of getting many diseases, including AD dementia.

Social determinants of health (SDOH) play a role in explaining health differences between groups of people. Education is the most studied SDOH in AD research studies, so we include it in your risk estimate.

**Race/ethnicity:** Evidence suggests some racial and ethnic groups may be at higher risk of AD dementia. For example, older adults who are of African Ancestry (Black, African American, African Caribbean, or African) may be more likely to get AD dementia than older adults who are White.

This difference may not actually be due to race. Other risk factors such as *APOE ε4*, family history, and social determinants of health may be the reason for differences between racial and ethnic groups.

### How can I find out my risk for AD dementia?

Current professional guidelines do not recommend returning genetic or biomarker results to healthy people. We are offering you the choice to receive your research results because some people in the MAP research studies have said they would like to see them.

If you choose, you can learn your research results and an estimate of your risk of getting AD dementia in the next 5 years based on your research results.

We will give you a risk estimate based on:

- Your age, gender, family history, and education
- 3 tests you already took as part of the Washington University MAP research studies:
    - Amyloid PET scan
    - Brain MRI
    - *APOE* genetic test results from your blood

We will give you a 5-year risk estimate based on your combined research results. We will also give you your individual research results. We are not currently returning tau results because the tests are very new and the evidence is less clear.

**If you choose to see your results, call our office to make an appointment.** That way, we can give you the report, explain the results, and answer any questions you have.

---

#### What you should know about research results

Make sure to talk to the study team before you share your results with doctors or if you want more information on clinical confirmation. Here's why:

There are different types of health testing. Your doctor orders a **clinical test** to help make healthcare decisions. Clinical tests are certified by the Food and Drug Administration (FDA). A **research test** may not have been certified by the FDA. Research tests may be done in different labs or using different machinery or methods. Research test results are not usually used to make health care decisions. Research tests are not usually placed in your medical record.

We are only returning research results that have scientific evidence about their relationship to AD dementia risk. Before you make any healthcare decisions, it's advised to confirm research test results with an FDA approved clinical test. There may be a cost and insurance implications if the test is clinically confirmed and put in your medical record.

### How can I decide if I want to know my risk?

We encourage you to think carefully about whether you want to know your research results and estimated risk of getting AD dementia. Once you know your results, you can't un-know them. For some people, their results may cause worry or anxiety.

#### Think about the pros and cons of finding out your risk.

**Here are some of the reasons people may want to know their research results. If you would like, you can make a checkmark by any that seem like good reasons for knowing:**

- ❑ Satisfy my curiosity
- ❑ Prepare myself and my family for the possibility of AD dementia
- ❑ Motivate me to lead a healthier lifestyle
- ❑ Arrange my personal affairs, such as retirement or financial planning
- ❑ Change my plans for the future or adjust the timing of when I will do things
- ❑ Treatments for AD dementia might become available
- ❑ I may be eligible for clinical trials
- ❑ Let my children know if they are at risk (genetic information only)
- ❑ My results could show that I am not at increased risk

**Here are some of the reasons people may not want to know their research results. If you would like, you can make a checkmark by any that seem like good reasons for not knowing:**

- ❑ I would be upset, anxious, or worry more if I am at increased risk for AD dementia
- ❑ My view of the future would change, or I would feel burdened to know
- ❑ There are currently no proven ways to prevent or cure AD dementia
- ❑ The results won't show if I definitely will get AD dementia
- ❑ If I share the results with a doctor, they could become part of my medical record
- ❑ The results may affect my ability to get insurance coverage
- ❑ If I share them, the results could change how other people interact with me
- ❑ My family and loved ones could be upset
- ❑ I might feel guilty if I found out I have a gene that could put my children at risk

We will not share your research results with your doctor(s). If you choose to share this information with your doctor(s), they may place it in your medical record. This may negatively affect future purchases of life or long-term care insurance. Make sure to discuss the implications before sharing this information with your doctor(s).

The Genetic Information Nondiscrimination Act (GINA) is a federal law that protects against genetic discrimination in health insurance and employment. GINA does not protect against discrimination in disability, life, or long-term care insurance. There are no laws currently that protect against discrimination based on brain imaging results.

#### You can ask yourself these questions to help you decide.

**If you would like, you can write your answers below or on a separate sheet of paper.**

- If I found out I have a higher or lower risk of getting AD dementia, what would that mean for me?

- How would I feel about information that is not certain or definite?

- What would I do with the information once I have it?

- How might the information change the way I feel or my views about other things in my life?

- Could the results affect getting insurance, such as long-term care coverage?

- What do my family or loved ones think? How might their behavior change?

### What can I do next?

#### Tell us if you want to see your research results and risk estimate.

After you have read this information, call us at 314-362-6535 if you would like to see your risk estimate based on your research results.

We will schedule an appointment where you will get your results report.

### What if I have questions?

If you have questions about getting your research results, contact the study team: **314-362-6535**.

### Resources

#### Find General AD information online or by calling:

- Alzheimer's Association, Greater Missouri Chapter — https://www.alz.org/greatermissouri Phone: 1-800-272-3900
- Missouri Association of Area Agencies on Aging — http://www.ma4web.org/
- AARP — https://www.aarp.org/aarp-foundation/

#### Learn about financial planning:

If you have concerns about long-term planning, visit these free online resources:

- Alzheimer Association Legal and Financial Planning: http://bit.ly/2OHmOoi
- US Government Consumer Finance: http://bit.ly/2OHCjwB
- US Government Consumer Finance Guide for Managing someone else's money: http://bit.ly/31F3Tjf

You can also contact an attorney. They can give you information on appointing a power of attorney for health decisions, trusts, assets, and long-term care planning.

- The Missouri Bar attorney search tool: https://bit.ly/2TY80E5

#### Talk to a licensed insurance broker:

If you have concerns about your insurance, we recommend you talk to a licensed health insurance broker.

#### Get more information about healthy eating and exercise:

Maintaining a healthy lifestyle, healthy eating, and getting exercise may help lower your risk of developing AD dementia. If you want more information on healthy eating and exercise, call your regular doctor (primary care physician, or PCP). They may refer you to a dietitian or social worker who can answer your questions and help develop healthy meal plans. This may be covered by your insurance. You can also look for local exercise classes to help you be active.

- Alzheimer Association "10 Ways to Love Your Brain:" https://bit.ly/3jSlBXZ
- National Institute on Aging "Healthy Eating:" https://bit.ly/3kWdXxl

### Example results report (Imaging)

KnightADRC Alzheimer's Disease Research Center | WASHINGTON UNIVERSITY ST. LOUIS

**These are research test results.** These results are not approved for clinical use by health care professionals, as the tests were performed for research purposes.

#### Research results prepared for [John Doe]

### We estimated your risk of developing early symptoms of AD dementia in the next 5 years.

##### Risk for people similar to you:

You were 78 when you had your MAP research scans.

Out of 100 men, 78 years of age, with a high school education, and with at least one parent who had AD dementia, **we estimate that:**

22 of them (22%) **will** develop AD dementia in the next 5 years.

That means the rest (78%) will **NOT** develop AD dementia in the next 5 years.

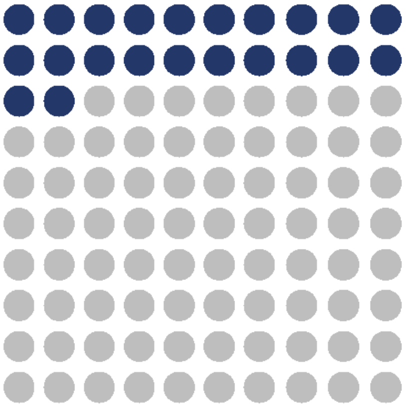

##### Your risk when we add your research results:

**When we consider the results of your brain imaging and blood tests, we estimate that:**

You have a **4%** chance of developing AD dementia in the next 5 years.

That means you have a **96%** chance of **NOT** developing AD dementia in the next 5 years.

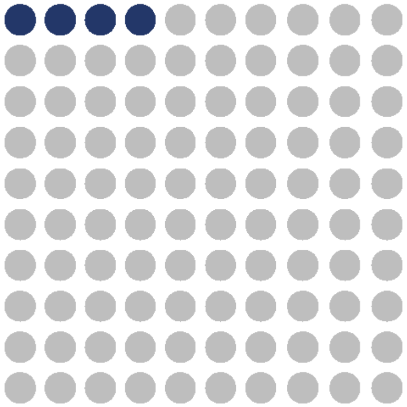

### How did you estimate my chance for AD dementia?

Your 5-year estimate of getting AD dementia considers how the results of your blood and brain imaging research tests interact with each other. This could make your combined risk of AD dementia somewhat different than your individual research results below. **Below you can learn more about the individual research results we used to get your risk estimate.**

#### *APOE* (apolipoprotein E) genetic test

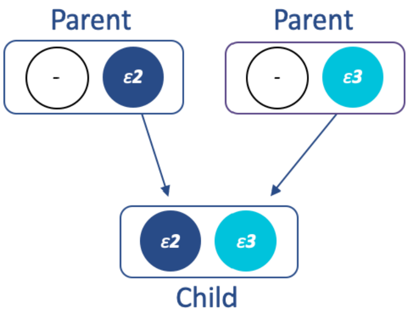

This section shows the results of the blood tests you had in the study. **We all inherit 2 copies of an *APOE* gene – one from each of our parents.** There are 3 different types of the *APOE* gene:

- ***APOE ε3* does not change** a person's risk of developing AD dementia. *ε3* is the most common type of *APOE*.
- ***APOE ε4* increases (raises)** a person's risk of developing AD dementia. *ε4* is the second most common type of APOE.
- ***APOE ε2* might decrease (lower)** a person's risk of developing AD dementia. *ε2* is the rarest type of *APOE*.

**Your APOE Genotype is ε2/ε3:** You have one copy of the *APOE* ***ε2*** gene and one copy of the *APOE* ***ε3*** gene. This result is associated with a decreased risk for AD dementia before we consider your other research results.

**This is a research genetic test result found through a research process.** This result is not approved for health care professional use in patient (clinical) care.

---

**Notes:**

**These are research test results.** These results are not approved for clinical use by health care professionals, as the tests were performed for research purposes.

#### Brain imaging tests

This section shows the results of the 2 types of brain imaging tests you had in the study: PET Amyloid scan and MRI scan.

##### PET Amyloid scan

This brain scan measures the amount of amyloid in a person's brain. Amyloid is a protein that is higher in the brains of people with AD dementia.

**Your PET Amyloid is *not elevated*.** This result shows an decreased risk for AD dementia, before we consider your other research results.

**This is a research PET scan result found through a research process.** This result is not approved for health care professional use in patient (clinical) care.

##### MRI scan

Your MRI gives a detailed scan of your brain. One of the things your MRI measures is a part of the brain called the hippocampus. The hippocampus plays a major role in memory. The hippocampus gets smaller as people age, but it is often substantially smaller in people with AD dementia. This measurement is called hippocampal volume.

**Your MRI result shows a *decreased risk* for AD dementia** on your estimated hippocampal volume, before we consider your other research results.

**This is a research MRI scan result found through a research process.** This result is not approved for health care professional use in patient (clinical) care.

---

Notes:

**These are research test results.** These results are not approved for clinical use by health care professionals, as the tests were performed for research purposes.

#### What should I know about research results?

Make sure to talk to the study team before you share your results with doctors or if you want more information on clinical confirmation. Here's why:

There are different types of health testing. Your doctor orders a **clinical test** to help make healthcare decisions. Clinical tests are certified by the Food and Drug Administration (FDA). A **research test** may not have been certified by the FDA. Research tests may be done in different labs or using different machinery or methods. Research test results are not usually used to make health care decisions. Research tests are not usually placed in your medical record.

We are only returning research results that have scientific evidence about their relationship to AD dementia risk . Before you make any healthcare decisions, it's advised to confirm research test results with an FDA approved clinical test. There may be a cost and insurance implications if the test is clinically confirmed and put in your medical record.

#### What if I have questions about these results later?

If you have questions about these research results after your appointment today, contact the study team:

Amy Oliver, MSW
314-362-6535

You can find more information and resources on AD dementia in the AD dementia brochure you received.

**These are research test results.** These results are not approved for clinical use by health care professionals, as the tests were performed for research purposes.

### Example results report (Plasma)

KnightADRC
Alzheimer's Disease Research Center
WASHINGTON UNIVERSITY ST. LOUIS

**These are research test results.** These results are not approved for clinical use by health care professionals, as the tests were performed for research purposes.

#### Research results prepared for [John Doe]

### We estimated your risk of developing <u>early symptoms</u> of AD dementia in the next 5 years.

##### Risk for people similar to you:

You were 81 when you had your MAP blood tests.

Out of 100 men, 81 years of age, with an education beyond high school, and without a parent who had AD dementia, **we estimate that:**

● 27 of them (27%) **will** develop AD dementia in the next 5 years.

● That means the rest (73%) will **NOT** develop AD dementia in the next 5 years.

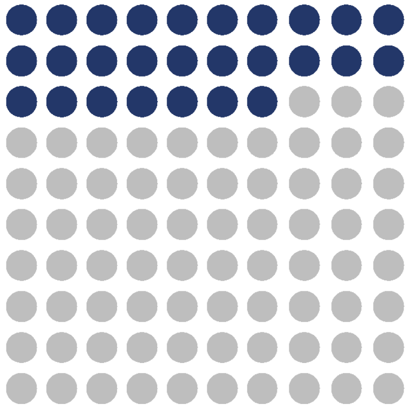

##### Your risk when we add your research results:

**When we consider the results of your blood tests, we estimate that:**

● You have a **36%** chance of developing AD dementia in the next 5 years.

● That means you have a **64%** chance of **NOT** developing AD dementia in the next 5 years.

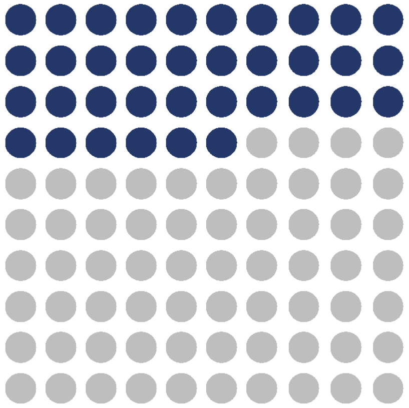

### How did you estimate my chance for AD dementia?

Your 5-year estimate of getting AD dementia considers how the results of your two blood tests, *APOE* genetic test and plasma amyloid test, interact with each other. This could make your combined risk of AD dementia somewhat different than your individual research results below. **Below you can learn more about the individual research results we used to get your risk estimate.**

#### *APOE* (apolipoprotein E) Genetic Test

This section shows the results of the blood tests you had in the study. **We all inherit 2 copies of an *APOE* gene – one from each of our parents.** There are 3 different types of the *APOE* gene:

- ***APOE ε3* does not change** a person's risk of developing AD dementia. *ε3* is the most common type of *APOE*.
- ***APOE ε4* increases (raises)** a person's risk of developing AD dementia. *ε4* is the second most common type of APOE.
- ***APOE ε2* might decrease (lower)** a person's risk of developing AD dementia. *ε2* is the rarest type of *APOE*.

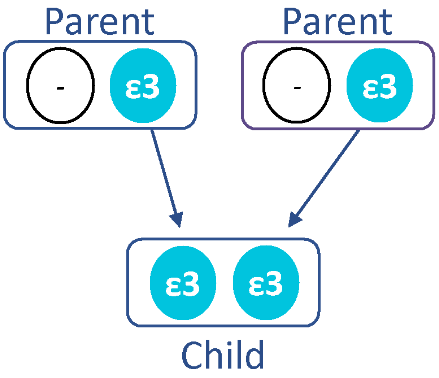

**Your *APOE* genotype is *ε3/ε3*:** You have one copy of the *APOE* ***ε3*** gene and one copy of the *APOE* ***ε3*** gene. This result is associated with *no change* in risk for AD dementia before we consider your other research results.

**This is a research genetic test result found through a research process.** This result is not approved for health care professional use in patient (clinical) care.

Notes:

**These are research test results.** These results are not approved for clinical use by health care professionals, as the tests were performed for research purposes.

### Amyloid Blood Test

This section shows the results of the blood test you had for this study.

#### Amyloid Blood Test

Amyloid, a protein that builds up in the brains of people with AD dementia, is also found in small amounts in the blood. The amount and types of amyloid found in the blood can tell us if amyloid is building up in the brain.

**Your amyloid blood test shows an *increased risk* for AD dementia**, before we consider your other research results.

**This is a research amyloid blood test result found through a research process.** This result is not approved for health care professional use in patient (clinical) care.

**These are research test results.** These results are not approved for clinical use by health care professionals, as the tests were performed for research purposes.

### Training manual for return of research results

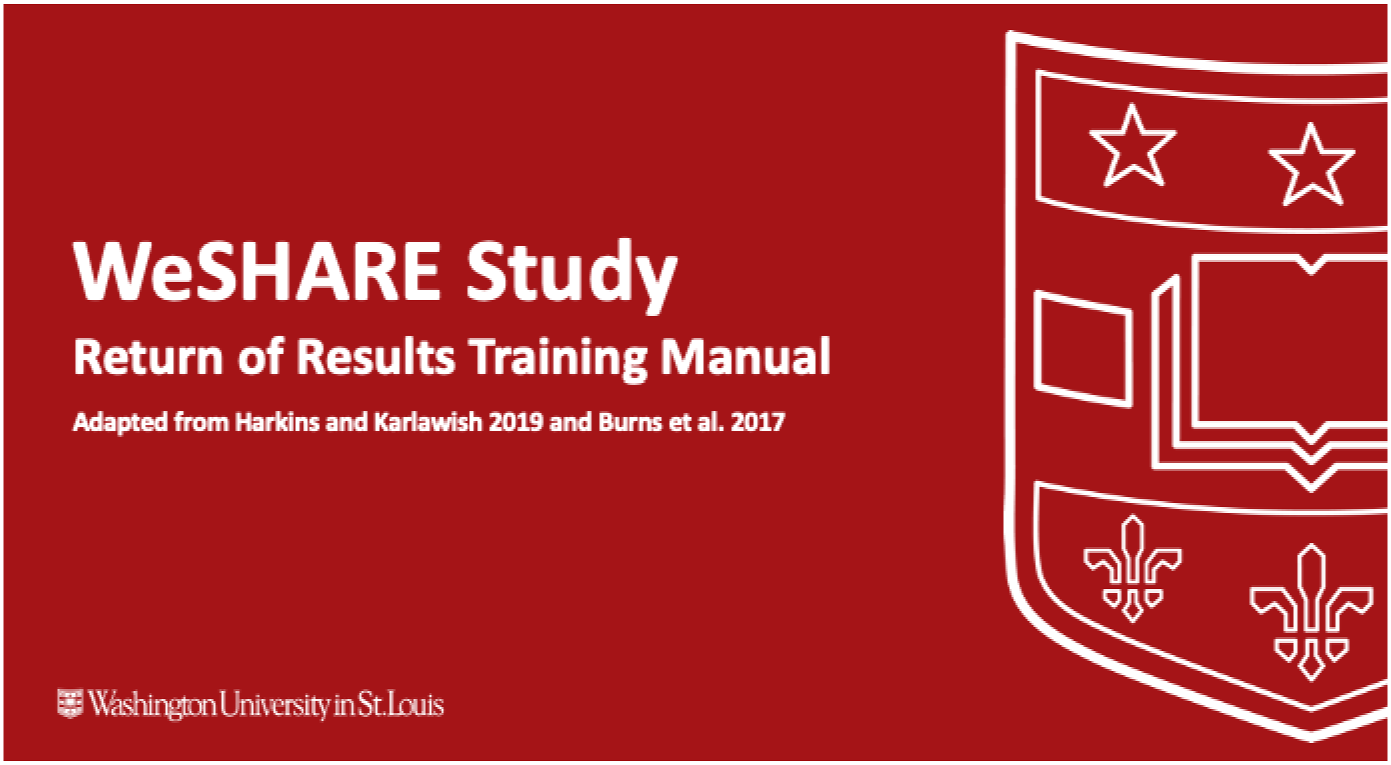

#### Table of Contents:

- Overview of Study Protocol
- General Guidelines for Return of Results
- Format of Reports
- Background Information
- Interpretation of Individual Test Results
- Case Examples
- Wrapping up the Session
- Frequently Asked Questions

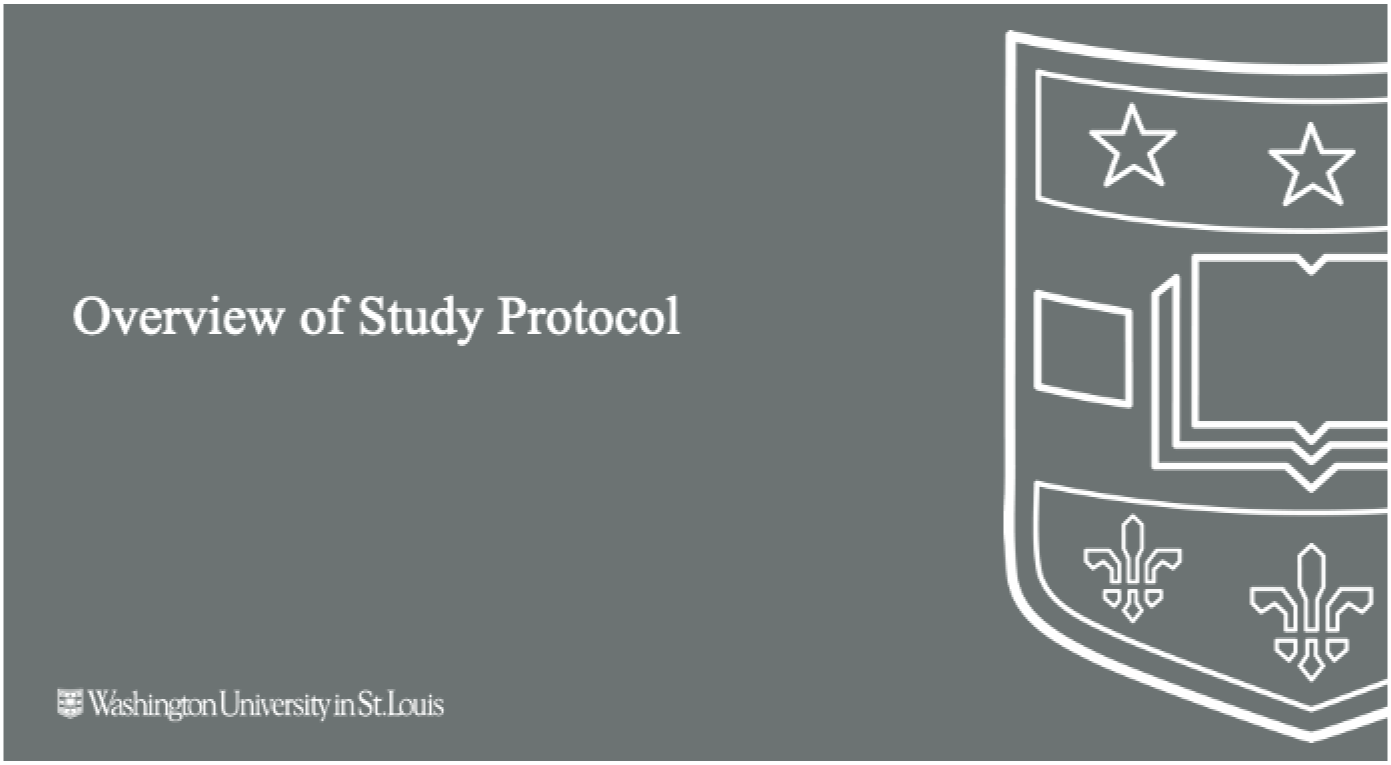
Overview of Study Protocol
Washington University in St. Louis

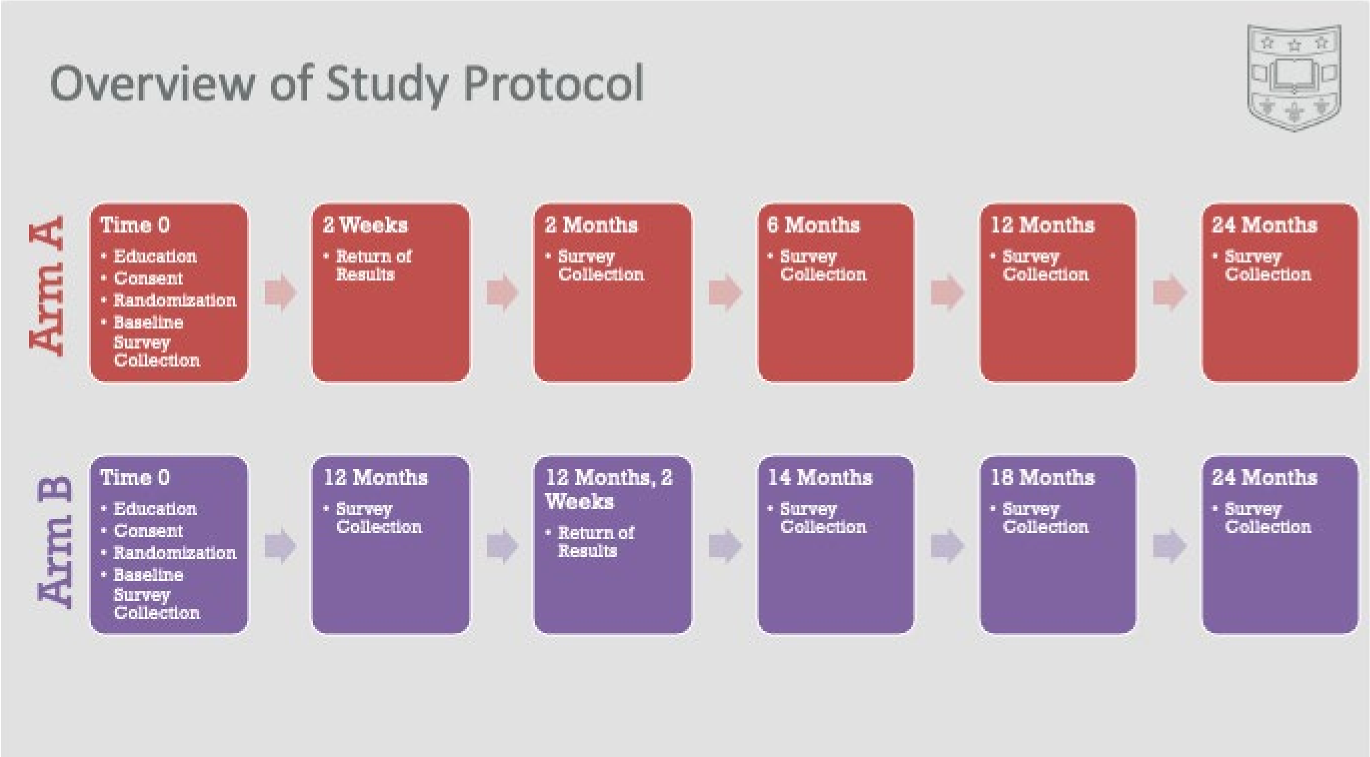
Overview of Study Protocol
Arm A
Time 0
• Education
• Consent
• Randomization
• Baseline Survey Collection
2 Weeks
• Return of Results
2 Months
• Survey Collection
6 Months
• Survey Collection
12 Months
• Survey Collection
24 Months
• Survey Collection
Arm B
Time 0
• Education
• Consent
• Randomization
• Baseline Survey Collection
12 Months
• Survey Collection
12 Months, 2 Weeks
• Return of Results
14 Months
• Survey Collection
18 Months
• Survey Collection
24 Months
• Survey Collection

### General Guidelines for Return of Results

Washington University in St. Louis

#### General Guidelines

**Emphasize that these are risk estimates only, not definitive, and based on what we know now.**

**Allow a support person to be present and leave adequate time for questions.**

**Monitor for signs of depression, anxiety, recent stressful news.**

**Avoid qualifying research results as "good" or "bad".**

- Researchers are not robots and if someone becomes emotional during a disclosure session – happy or sad – then it is appropriate to respond to them and acknowledge their feelings and be responsive to them. We just want to avoid framing things in advance for them where possible.

**The "teach back method" is an evidence-based practice that can be used during participant education to assess patient understanding (more information on slide 53).**

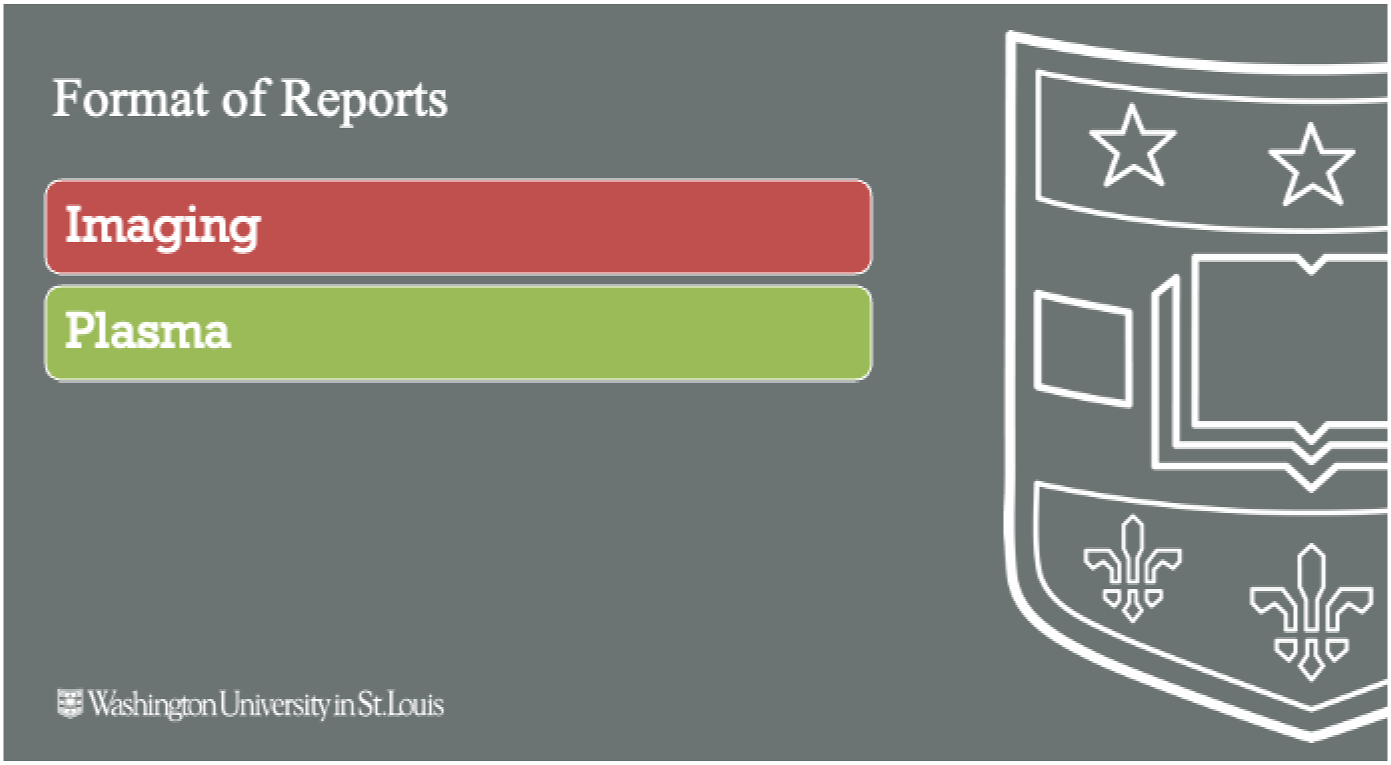
Format of Reports
Imaging
Plasma
Washington University in St. Louis

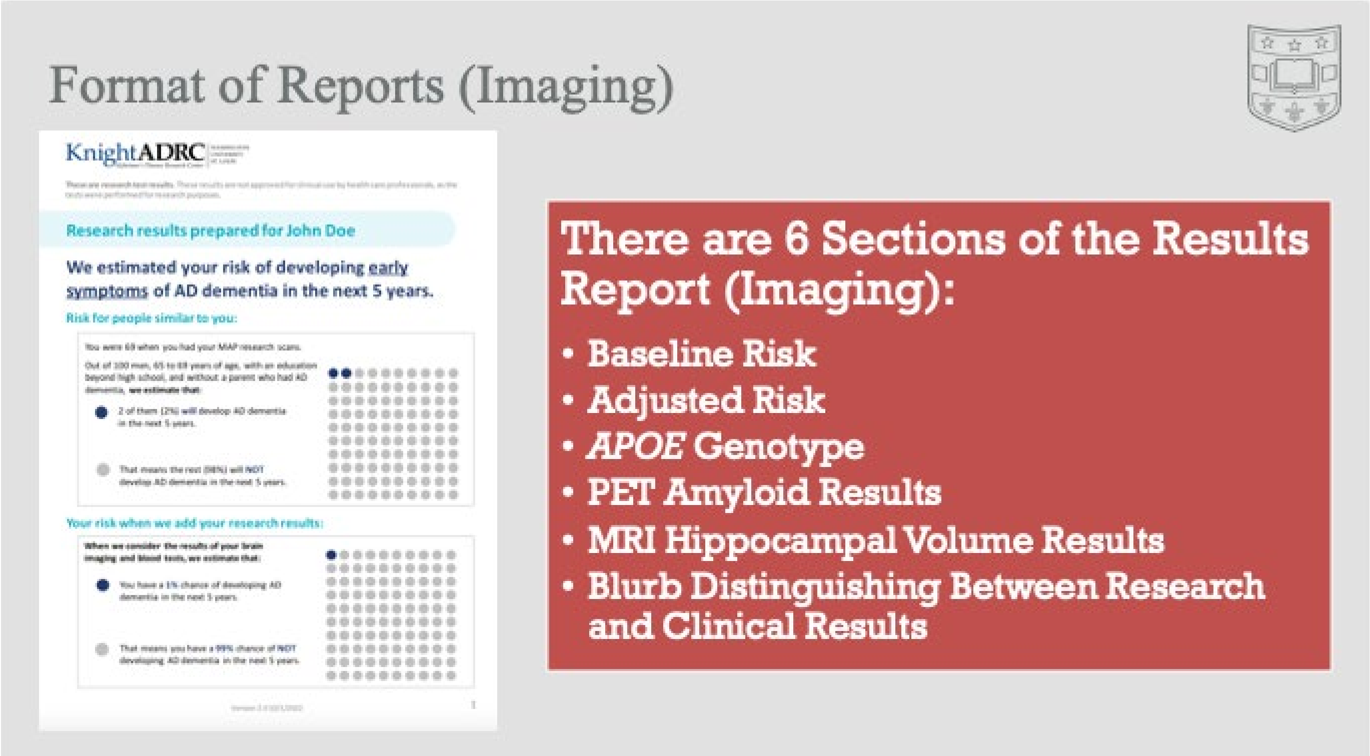
Format of Reports (Imaging)
KnightADRC
Research results prepared for John Doe
We estimated your risk of developing early symptoms of AD dementia in the next 5 years.
Risk for people similar to you:
Your risk when we add your research results:
There are 6 Sections of the Results Report (Imaging):
• Baseline Risk
• Adjusted Risk
• APOE Genotype
• PET Amyloid Results
• MRI Hippocampal Volume Results
• Blurb Distinguishing Between Research and Clinical Results

#### Format of Reports (Plasma)

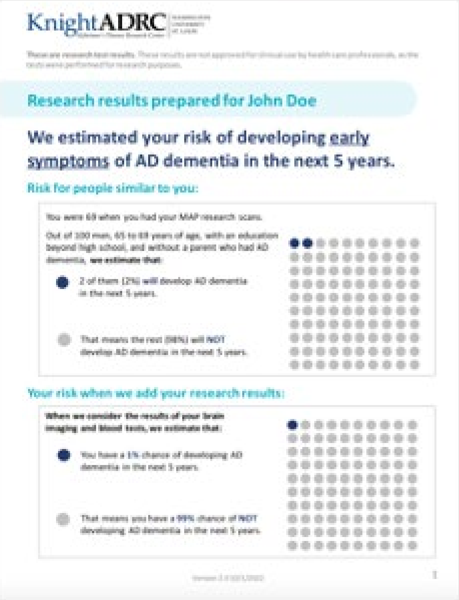
KnightADRC

Research results prepared for John Doe

We estimated your risk of developing early symptoms of AD dementia in the next 5 years.

Risk for people similar to you:

You were 69 when you had your MAP research scans.

Out of 100 men, 65 to 69 years of age, with an education beyond high school, and without a parent who had AD dementia, **we estimate that:**

- 2 of them (2%) will develop AD dementia in the next 5 years.
- That means the rest (98%) will NOT develop AD dementia in the next 5 years.

Your risk when we add your research results:

**When we consider the results of your brain imaging and blood tests, we estimate that:**

- You have a 1% chance of developing AD dementia in the next 5 years.
- That means you have a 99% chance of NOT developing AD dementia in the next 5 years.

##### There are 5 Sections of the Results Report (Plasma):

- Baseline Risk
- Adjusted Risk
- *APOE* Genotype
- Plasma Amyloid Results
- Blurb Distinguishing Between Research and Clinical Results

#### Background Information

- Baseline Risk
- Adjusted Risk
- *APOE* Genotype
- Amyloid (PET)
- Hippocampal Volume (MRI)
- Amyloid (Plasma)

Washington University in St. Louis

#### Background: Baseline Risk

1. First, we calculated a risk estimate of developing early signs of AD in the next 5 years, based on demographics (prior to looking at any research results).

2. If we looked at 100 people who were [X demographics] we estimate [X] would develop early symptoms of AD in the next five years.

You were 69 when you had your MAP research scans.

Out of 100 men, 65 to 69 years of age, with an education beyond high school, and without a parent who had AD dementia, **we estimate that:**

- 2 of them (2%) **will** develop AD dementia in the next 5 years.
- That means the rest (98%) will **NOT** develop AD dementia in the next 5 years.

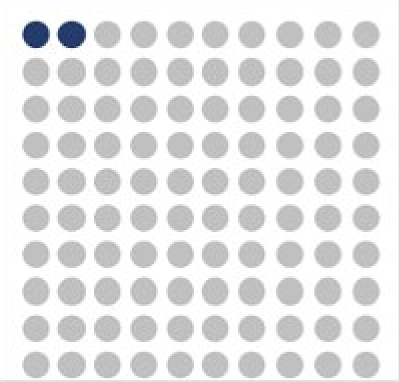

#### Background: Adjusted Risk

1. An adjusted risk estimate is calculated using demographics as well as research results from the Memory and Aging Project (MAP) study.

2. The research results interact with each other in ways that could make a combined risk of AD dementia different than risk from individual research results.

3. For example, overall risk estimate could suggest a decreased risk of AD dementia even though some individual research results suggest an increased risk.

- *Analogy: If a meteorologist only looked at the humidity level without looking at the air temperature or wind speed, the estimated chance of rain could be different than when all the data points are included in the prediction model.*

**When we consider the results of your brain imaging and blood tests, we estimate that:**

- You have a 1% chance of developing AD dementia in the next 5 years.
- That means you have a 99% chance of NOT developing AD dementia in the next 5 years.

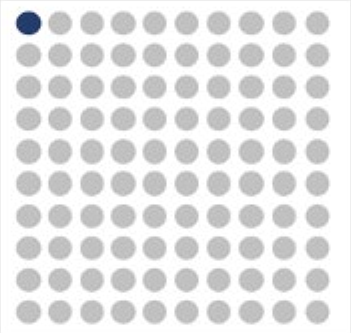

#### Background: *APOE* Genotype

1. Everyone has two copies of the *APOE* gene, one that they inherit from their biological mother and one that they inherit from their biological father.
2. There are three different alleles (versions) of the *APOE* gene, that are associated with different risks for developing AD:
   - **ε3 allele– neutral risk**
     - Most common form:
       - ~ 60% of people have two copies of the ε3 allele.
   - **ε4 allele–increases risk**
     - Second most common form:
       - ~ 25% of people have one copy of the ε4 allele.
       - ~2-3% of people have two copies of the ε4 allele.
     - Increased Risk:
       - Those who inherit one copy of the ε4 form have about three times the risk of developing Alzheimer's compared with those with two copies of the ε3 form.
       - Those who inherit two copies of the ε4 form have about eight-twelve times the risk of developing Alzheimer's compared with those with two copies of the ε3 form.
   - **ε2 allele–decreases risk**
     - Least common form:
       - ~10% of people have one copy of the ε2 allele.
       - ~0.5% of people have two copies of the ε2 allele.
3. Penetrance
   - The ε4 allele is a risk factor, but is not necessary (many people without an ε4 allele will develop AD dementia) nor sufficient (many people with an ε4 allele will NOT develop AD dementia) to develop AD.
4. The reason *APOE* ε4 increases Alzheimer's risk is not well understood. The APOE protein helps carry cholesterol and other types of fat in the bloodstream. Recent studies suggest that problems with brain cells' ability to process fats, or lipids, may play a key role in Alzheimer's and related diseases.
5. Most of the research to date associating *APOE* ε4 with increased risk of Alzheimer's has studied White individuals. Studies of this association in Black and Hispanic populations have had inconsistent results.

#### Background: Amyloid (PET)

1. Having elevated levels of beta-amyloid plaques in the brain is a hallmark trait of AD dementia.
2. Amyloid PET is a snapshot of the amount of beta-amyloid plaques in the brain.
3. There is a standard cutoff that we use to determine if amyloid is elevated or not:
   - **Elevated Amyloid:** the level of amyloid is above this cutoff
     - All people with AD dementia have elevated amyloid, but not all people with elevated amyloid get AD dementia.
     - This means you could live the rest of your life with elevated amyloid and never develop signs and symptoms of AD dementia.
   - **Not Elevated Amyloid:** the level of amyloid is below this cutoff.
     - Amyloid level could become elevated in the future.
     - There is still a chance of developing AD dementia in future.

#### Background: Hippocampal Volume (MRI)

1. Magnetic resonance imaging (MRI) can be used to measure different structures of the brain, including the hippocampus.
2. Hippocampal volume often decreases with age, and is known to be smaller in individuals with AD.
3. We created a cutoff for increased hippocampal volumes.
   – We expect 50% of people in this study will get hippocampal volumes above the cutoff and 50% will get hippocampal volumes below the cutoff.
4. **Above average hippocampal volumes:** above average for healthy people 60 or older.
   – Associated with a decreased risk for developing AD
5. **Below average hippocampal volumes:** below average for healthy people 60 or older.
   – Associated with an increased risk for developing AD
      - This does not mean that the participant will definitely get AD dementia in the future.
      - This does not necessarily mean that the participant's hippocampal volume has decreased from prior scans, it just means that it is smaller than the cutoff.

#### Background: Amyloid (Plasma)

1. Traditionally we have used PET scans to measure Amyloid levels in the brain. Now there is a blood test where we can see if you are likely to have the presence or absence of amyloid plaques in the brain.
   – The test measures amyloid-beta 42/40 ratio (Aβ 42/40) using blood mass spectrometry technology.
   – Both PET and plasma can measure the amount of amyloid on the brain. The plasma test uses newer technology that is not quite as accurate as PET. This is why when imaging if available, we use that. The new blood test allows us to return results to participants who do not/cannot have imaging.
2. **Elevated Amyloid:** the level of amyloid is above this cutoff.
   – All people with AD dementia have elevated amyloid, but not all people with elevated amyloid get AD dementia.
   – This means you could live the rest of your life with elevated amyloid and never develop signs and symptoms of AD dementia.
3. **Not Elevated Amyloid:** the level of amyloid is below this cutoff.
   – Amyloid level could become elevated in the future.
   – There is still a chance of developing AD dementia in future.

### Interpretations of Individual Test Results

*APOE* **Genotype**

**Amyloid (PET)**

**Hippocampal Volume (MRI)**

**Amyloid (Plasma)**

Washington University in St. Louis

---

##### How did you estimate my chance for AD dementia?

Your 5-year estimate of getting AD dementia considers how the results of your blood and brain imaging research tests interact with each other. This could make your combined risk of AD dementia somewhat different than your individual research results below. **Below you can learn more about the individual research results we used to get your risk estimate.**

###### *APOE* (apolipoprotein E) genetic test

This section shows the results of the blood tests you had in the study. **We all inherit 2 copies of an *APOE* gene – one from each of our parents.** There are 3 different types of the *APOE* gene:

- ***APOE* ε3 does not change** a person's risk of developing AD dementia. ε3 is the most common type of *APOE*.
- ***APOE* ε4 increases (raises)** a person's risk of developing AD dementia. ε4 is the second most common type of *APOE*.
- ***APOE* ε2 might decrease (lower)** a person's risk of developing AD dementia. ε2 is the rarest type of *APOE*.

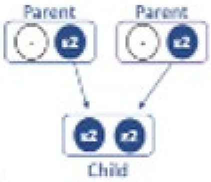

Your *APOE* Genotype is ε2/ε2: You have one copy of the *APOE* ε2 gene and one copy of the *APOE* ε2 gene. This result is associated with a decreased risk for AD dementia before we consider your other research results.

**This is a research genetic test result found through a research process.** This result is not approved for health care professional use in patient (clinical) care.

Notes:

#### *APOE* Genotype: ε2/ε2

- You may remember having your blood drawn as part of the memory and aging project study. One of the tests that they looked at is a genetic test, looking at a gene called *APOE*.
- Everyone has two copies of the *APOE* gene, one they they inherit from their biological mother and one that they inherit from their biological father.
  - The most common is the ε3 type, which is neutral (does not increase or decrease) risk for developing AD.
  - The second most common is the ε4 type which is known to increase risk for developing AD.
  - The least common is the ε2 type which is known to be protective or decrease risk for developing AD.
- [Point to figure]: In this figure you are the child. You inherited two copies of the rare ε2 type, which is associated with a decreased in risk in AD dementia before looking at any of your other research results.
- Any questions?

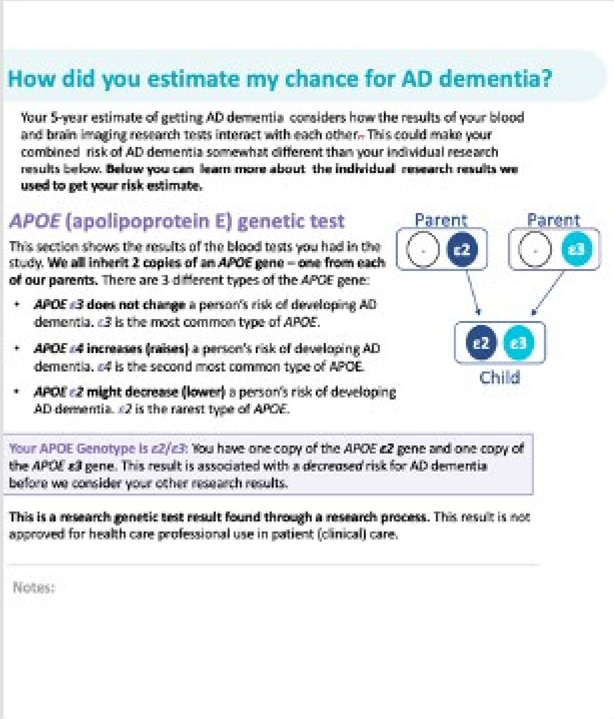

#### How did you estimate my chance for AD dementia?

Your 5-year estimate of getting AD dementia considers how the results of your blood and brain imaging research tests interact with each other. This could make your combined risk of AD dementia somewhat different than your individual research results below. **Below you can learn more about the individual research results we used to get your risk estimate.**

##### *APOE* (apolipoprotein E) genetic test

This section shows the results of the blood tests you had in the study. **We all inherit 2 copies of an *APOE* gene – one from each of our parents.** There are 3 different types of the *APOE* gene:

- ***APOE* ε3 does not change** a person's risk of developing AD dementia. *ε3* is the most common type of *APOE*.
- ***APOE* ε4 increases (raises)** a person's risk of developing AD dementia. ε4 is the second most common type of APOE.
- ***APOE* ε2 might decrease (lower)** a person's risk of developing AD dementia. *ε2* is the rarest type of *APOE*.

Your APOE Genotype is *ε2/ε3*: You have one copy of the *APOE* **ε2** gene and one copy of the *APOE* **ε3** gene. This result is associated with a *decreased* risk for AD dementia before we consider your other research results.

**This is a research genetic test result found through a research process.** This result is not approved for health care professional use in patient (clinical) care.

Notes:

### *APOE* Genotype: ε2/ε3

- You may remember having your blood drawn as part of the memory and aging project study. One of the tests that they looked at is a genetic test, looking at a gene called *APOE*.
- Everyone has two copies of the *APOE* gene, one they they inherit from their biological mother and one that they inherit from their biological father.
  - The most common is the ε3 type, which is neutral (does not increase or decrease) risk for developing AD.
  - The second most common is the ε4 type which is known to increase risk for developing AD.
  - The least common is the ε2 type which is known to be protective or decrease risk for developing AD.
- [Point to figure]: In this figure, you are the child. You inherited one copy of the protective ε2 type, and one copy of the neutral ε3 type, which is associated with an overall decreased in risk in AD dementia before looking at any of your other research results.
- Any questions?

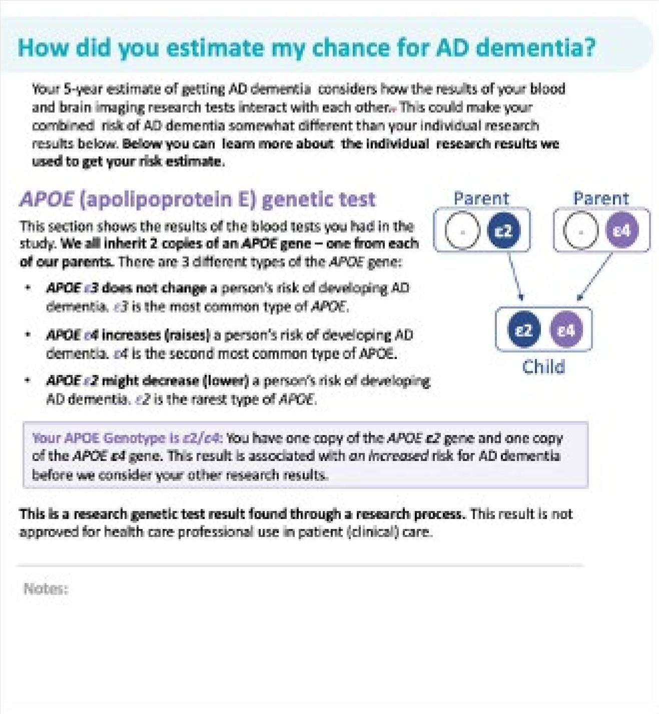

#### How did you estimate my chance for AD dementia?

Your 5-year estimate of getting AD dementia considers how the results of your blood and brain imaging research tests interact with each other. This could make your combined risk of AD dementia somewhat different than your individual research results below. **Below you can learn more about the individual research results we used to get your risk estimate.**

##### *APOE* (apolipoprotein E) genetic test

This section shows the results of the blood tests you had in the study. **We all inherit 2 copies of an *APOE* gene – one from each of our parents.** There are 3 different types of the *APOE* gene:

- ***APOE* ε3 does not change** a person's risk of developing AD dementia. *ε3* is the most common type of *APOE*.
- ***APOE* ε4 increases (raises)** a person's risk of developing AD dementia. ε4 is the second most common type of APOE.
- ***APOE* ε2 might decrease (lower)** a person's risk of developing AD dementia. *ε2* is the rarest type of *APOE*.

Your APOE Genotype is *ε2/ε4*: You have one copy of the *APOE* **ε2** gene and one copy of the *APOE* **ε4** gene. This result is associated with *an increased* risk for AD dementia before we consider your other research results.

**This is a research genetic test result found through a research process.** This result is not approved for health care professional use in patient (clinical) care.

Notes:

### *APOE* Genotype: ε2/ε4

- You may remember having your blood drawn as part of the memory and aging project study. One of the tests that they looked at is a genetic test, looking at a gene called *APOE*.
- Everyone has two copies of the *APOE* gene, one they they inherit from their biological mother and one that they inherit from their biological father.
  - The most common is the ε3 type, which is neutral (does not increase or decrease) risk for developing AD.
  - The second most common is the ε4 type which is known to increase risk for developing AD.
  - The least common is the ε2 type which is known to be protective or decrease risk for developing AD.
- [Point to figure]: In this figure, you are the child. You inherited one copy of the ε2 type and one copy of the ε4 type.
- This combination is less well understood, because ε2 decreases risk, where ε4 increases risk. It is thought that there may be a slight increase in risk (likely more than if someone had ε3/ε3, but less risk than ε3/ε4.
- ~ 30% of the general population has at least one copy of the ε4 gene, so it is possible to have one or even two copies of ε4 *APOE* gene and never develop symptoms of AD.
- Any questions?

#### How did you estimate my chance for AD dementia?

Your 5-year estimate of getting AD dementia considers how the results of your blood and brain imaging research tests interact with each other. This could make your combined risk of AD dementia somewhat different than your individual research results below. **Below you can learn more about the individual research results we used to get your risk estimate.**

##### *APOE* (apolipoprotein E) genetic test

This section shows the results of the blood tests you had in the study. **We all inherit 2 copies of an *APOE* gene – one from each of our parents.** There are 3 different types of the *APOE* gene:

- ***APOE* ε3 does not change** a person's risk of developing AD dementia. ε3 is the most common type of *APOE*.
- ***APOE* ε4 increases (raises)** a person's risk of developing AD dementia. ε4 is the second most common type of APOE.
- ***APOE* ε2 might decrease (lower)** a person's risk of developing AD dementia. ε2 is the rarest type of *APOE*.

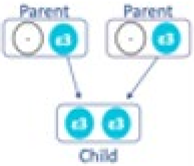

Your APOE Genotype is ε3/ε3: You have two copies of the *APOE ε3* gene. This result is associated with *no change* in risk for AD dementia before we consider your other research results.

**This is a research genetic test result found through a research process.** This result is not approved for health care professional use in patient (clinical) care.

Notes:

### *APOE* Genotype: ε3/ε3

- You may remember having your blood drawn as part of the memory and aging project study. One of the tests that they looked at is a genetic test, looking at a gene called *APOE*.
- Everyone has two copies of the *APOE* gene, one they they inherit from their biological mother and one that they inherit from their biological father.
  - The most common is the ε3 type, which is neutral (does not increase or decrease) risk for developing AD.
  - The second most common is the ε4 type which is known to increase risk for developing AD.
  - The least common is the ε2 type which is known to be protective or decrease risk for developing AD.
- [Point to figure]: In this figure you are the child. You inherited two copies of the common ε3 type, which is associated which does not change your risk of developing AD.
- Any questions?

#### How did you estimate my chance for AD dementia?

Your 5-year estimate of getting AD dementia considers how the results of your blood and brain imaging research tests interact with each other. This could make your combined risk of AD dementia somewhat different than your individual research results below. **Below you can learn more about the individual research results we used to get your risk estimate.**

##### *APOE* (apolipoprotein E) genetic test

This section shows the results of the blood tests you had in the study. **We all inherit 2 copies of an *APOE* gene – one from each of our parents.** There are 3 different types of the *APOE* gene:

- ***APOE* ε3 does not change** a person's risk of developing AD dementia. ε3 is the most common type of *APOE*.
- ***APOE* ε4 increases (raises)** a person's risk of developing AD dementia. ε4 is the second most common type of APOE.
- ***APOE* ε2 might decrease (lower)** a person's risk of developing AD dementia. ε2 is the rarest type of *APOE*.

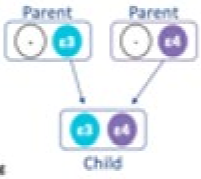

Your APOE Genotype is ε3/ε4: You have one copy of the *APOE ε3* gene and one copy of the *APOE ε4* gene. This result is associated with *an increased* risk for AD dementia before we consider your other research results.

**This is a research genetic test result found through a research process.** This result is not approved for health care professional use in patient (clinical) care.

Notes:

### *APOE* Genotype: ε3/ε4

- You may remember having your blood drawn as part of the memory and aging project study. One of the tests that they looked at is a genetic test, looking at a gene called *APOE*.
- Everyone has two copies of the *APOE* gene, one they they inherit from their biological mother and one that they inherit from their biological father.
  - The most common is the ε3 type, which is neutral (does not increase or decrease) risk for developing AD.
  - The second most common is the ε4 type which is known to increase risk for developing AD.
  - The least common is the ε2 type which is known to be protective or decrease risk for developing AD.
- [Point to figure]: In this figure, you are the child. You inherited one copy of the neutral ε3 type and one copy of the ε4 type which is associated with an overall increased in risk in AD dementia before looking at any of your other research results.
- Those who inherit one copy of the ε4 form have about three times the risk of developing Alzheimer's compared with those with two copies of the ε3 form.
- ~30% of the general population has at least one copy of the ε4 gene, so it is possible to have one or even two copies of ε4 *APOE* gene and never develop symptoms of AD.
- Any questions?

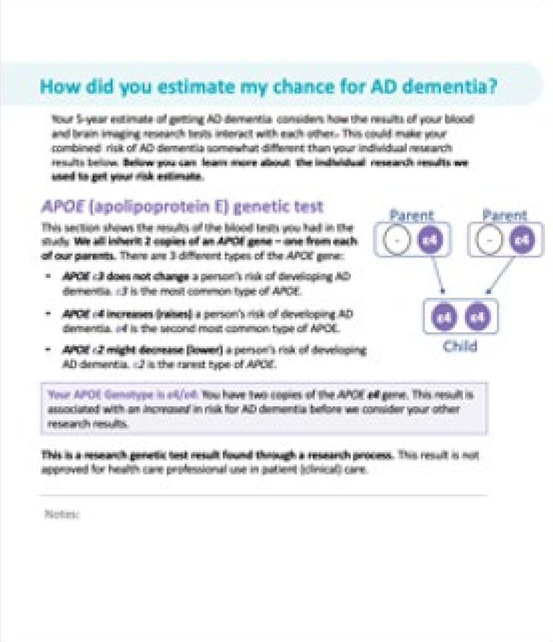

#### How did you estimate my chance for AD dementia?

Your 5-year estimate of getting AD dementia considers how the results of your blood and brain imaging research tests interact with each other. This could make your combined risk of AD dementia somewhat different than your individual research results below. **Below you can learn more about the individual research results we used to get your risk estimate.**

##### *APOE* (apolipoprotein E) genetic test

This section shows the results of the blood tests you had in the study. **We all inherit 2 copies of an *APOE* gene – one from each of our parents.** There are 3 different types of the *APOE* gene:

- ***APOE* ε3 does not change** a person's risk of developing AD dementia. ε3 is the most common type of *APOE*.
- ***APOE* ε4 increases (raises)** a person's risk of developing AD dementia. ε4 is the second most common type of *APOE*.
- ***APOE* ε2 might decrease (lower)** a person's risk of developing AD dementia. ε2 is the rarest type of *APOE*.

Your APOE Genotype is ε4/ε4: You have two copies of the *APOE* **ε4** gene. This result is associated with an *increased* in risk for AD dementia before we consider your other research results.

**This is a research genetic test result found through a research process.** This result is not approved for health care professional use in patient (clinical) care.

Notes:

### *APOE* Genotype: ε4/ε4

- You may remember having your blood drawn as part of the memory and aging project study. One of the tests that they looked at is a genetic test, looking at a gene called *APOE*.
- Everyone has two copies of the *APOE* gene, one they they inherit from their biological mother and one that they inherit from their biological father.
  - The most common is the ε3 type, which is neutral (does not increase or decrease) risk for developing AD.
  - The second most common is the ε4 type which is known to increase risk for developing AD.
  - The least common is the ε2 type which is known to be protective or decrease risk for developing AD.
- [Point to figure]: In this figure, you are the child. You inherited two copies of the ε4 type which is associated with an overall increased in risk in AD dementia before looking at any of your other research results.
- Those who inherit two copies of the ε4 form have about eight-twelve times the risk of developing Alzheimer's compared with those with two copies of the ε3 form
- ~ 30% of the general population has at least one copy of the ε4 gene, so it is possible to have one or even two copies of ε4 *APOE* gene and never develop symptoms of AD.
- Any questions?

### Amyloid (PET): Elevated

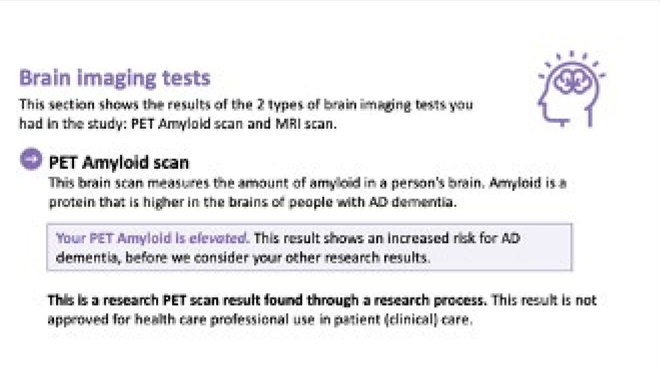

#### Brain imaging tests

This section shows the results of the 2 types of brain imaging tests you had in the study: PET Amyloid scan and MRI scan.

##### PET Amyloid scan

This brain scan measures the amount of amyloid in a person's brain. Amyloid is a protein that is higher in the brains of people with AD dementia.

Your PET Amyloid is *elevated*. This result shows an increased risk for AD dementia, before we consider your other research results.

**This is a research PET scan result found through a research process.** This result is not approved for health care professional use in patient (clinical) care.

- You may remember having a PET scan as part of the memory and aging project.
- One of the scans they did was looking at a protein in the brain called amyloid.
- Amyloid can build up in the brain as people age, but we know that there tends to be greater levels of amyloid in the brains of people with AD dementia.
- When looking at your PET scan data, we saw that your amyloid levels were elevated, indicating an increased risk for developing AD.
- Elevated amyloid does not guarantee that you will develop AD, but it is associated with increased risk before we consider any of your other research results.
- Questions?

#### Amyloid (PET): Not Elevated

**Brain imaging tests**

This section shows the results of the 2 types of brain imaging tests you had in the study: PET Amyloid scan and MRI scan.

**PET Amyloid scan**

This brain scan measures the amount of amyloid in a person's brain. Amyloid is a protein that is higher in the brains of people with AD dementia.

> Your PET Amyloid is *not elevated*. This result shows a *decreased risk* for AD dementia, before we consider your other research results.

**This is a research PET scan result found through a research process.** This result is not approved for health care professional use in patient (clinical) care.

- You may remember having a PET scan as part of the memory and aging project.
- One of the scans they did was looking at a protein in the brain called amyloid.
- Amyloid can build up in the brain as people age, but we know that there tends to be greater levels of amyloid in the brains of people with AD dementia.
- When looking at your PET scan data, we saw that your amyloid levels were NOT elevated, indicating a decreased risk for developing AD before considering any of your other research results.
- It is possible amyloid levels could increase in the future.
- Questions?

#### Hippocampal Volume (MRI): Decreased Volume

**MRI scan**

Your MRI gives a detailed scan of your brain. One of the things your MRI measures is a part of the brain called the hippocampus. The hippocampus plays a major role in memory. The hippocampus gets smaller as people age, but it is often substantially smaller in people with AD dementia. This measurement is called hippocampal volume.

> Your MRI result shows an *increased risk* for AD dementia based on your estimated hippocampal volume, before we consider your other research results.

**This is a research MRI scan result found through a research process.** This result is not approved for health care professional use in patient (clinical) care.

Notes:

- You may remember having an MRI scan as part of the memory and aging project.
- One of the things that they look at on the MRI images is called hippocampal volume (HV).
- The hippocampus plays an important role in memory. We know that hippocampal volume size decreases as people age, but tends to be smaller in individuals with AD.
- When looking at your MRI scans, we saw that your HV was decreased, indicating an increased risk for developing AD.
- This means that your HV was smaller than a certain cutoff, NOT that your HV has necessarily decreased from prior scans.
- Decreased HV does not guarantee that you will develop AD, but it is associated with increased risk before we consider any of your other research results.
- Questions?

### Hippocampal Volume (MRI): Typical Volume

**MRI scan**

Your MRI gives a detailed scan of your brain. One of the things your MRI measures is a part of the brain called the hippocampus. The hippocampus plays a major role in memory. The hippocampus gets smaller as people age, but it is often substantially smaller in people with AD dementia. This measurement is called hippocampal volume.

Your MRI result shows a *decreased risk* for AD dementia based on your estimated hippocampal volume, before we consider your other research results.

**This is a research MRI scan result found through a research process.** This result is not approved for health care professional use in patient (clinical) care.

Notes:

- You may remember having an MRI scan as part of the memory and aging project.
- One of the things that they look at on the MRI images is called hippocampal volume (HV).
- The hippocampus plays an important role in memory.
- We know that hippocampal volume size decreases as people age, but tends to be smaller in individuals with AD.
- When looking at your MRI scans, we saw that your HV was NOT decreased, indicating a decreased risk for developing AD before we consider any of your other research results.
- Questions?

### Amyloid (Plasma): Elevated

#### Amyloid Blood Test

This section shows the results of the blood test you had for this study.

**Amyloid Blood Test**

Amyloid, a protein that builds up in the brains of people with AD dementia, is also found in small amounts in the blood. The amount and types of amyloid found in the blood can tell us if amyloid is building up in the brain.

Your amyloid blood test shows an *increased risk* for AD dementia, before we consider your other research results.

**This is a research amyloid blood test result found through a research process.** This result is not approved for health care professional use in patient (clinical) care.

- One of the other things that they were looking at in your blood a protein in the brain called amyloid.
- Amyloid can build up in the brain as people age, but we know that there tends to be greater levels of amyloid in the brains of people with AD dementia.
- When looking at your data, we saw that your amyloid levels were elevated, indicating an increased risk for developing AD.
- Elevated amyloid does not guarantee that you will develop AD, but it is associated with increased risk before we consider any of your other research results.
- Questions?

#### Amyloid Blood Test

This section shows the results of the blood test you had for this study.

##### Amyloid Blood Test

Amyloid, a protein that builds up in the brains of people with AD dementia, is also found in small amounts in the blood. The amount and types of amyloid found in the blood can tell us if amyloid is building up in the brain.

> Your amyloid blood test shows a decreased *risk* for AD dementia, before we consider your other research results.

**This is a research amyloid blood test result found through a research process.** This result is not approved for health care professional use in patient (clinical) care.

### Amyloid (Plasma): Not Elevated

- One of the other things that they were looking at in your blood a protein in the brain called amyloid.
- Amyloid can build up in the brain as people age, but we know that there tends to be greater levels of amyloid in the brains of people with AD dementia.
- When looking at your data, we saw that your amyloid levels were NOT elevated, indicating an decreased risk for developing AD before we consider any of your other research results.
- It is possible amyloid levels could increase in the future.
- Questions?

### Examples

The research results interact with each other in ways that could make a combined risk of AD dementia different than risk from individual research results. Please see three examples of participant reports on the following slides.

Washington University in St. Louis

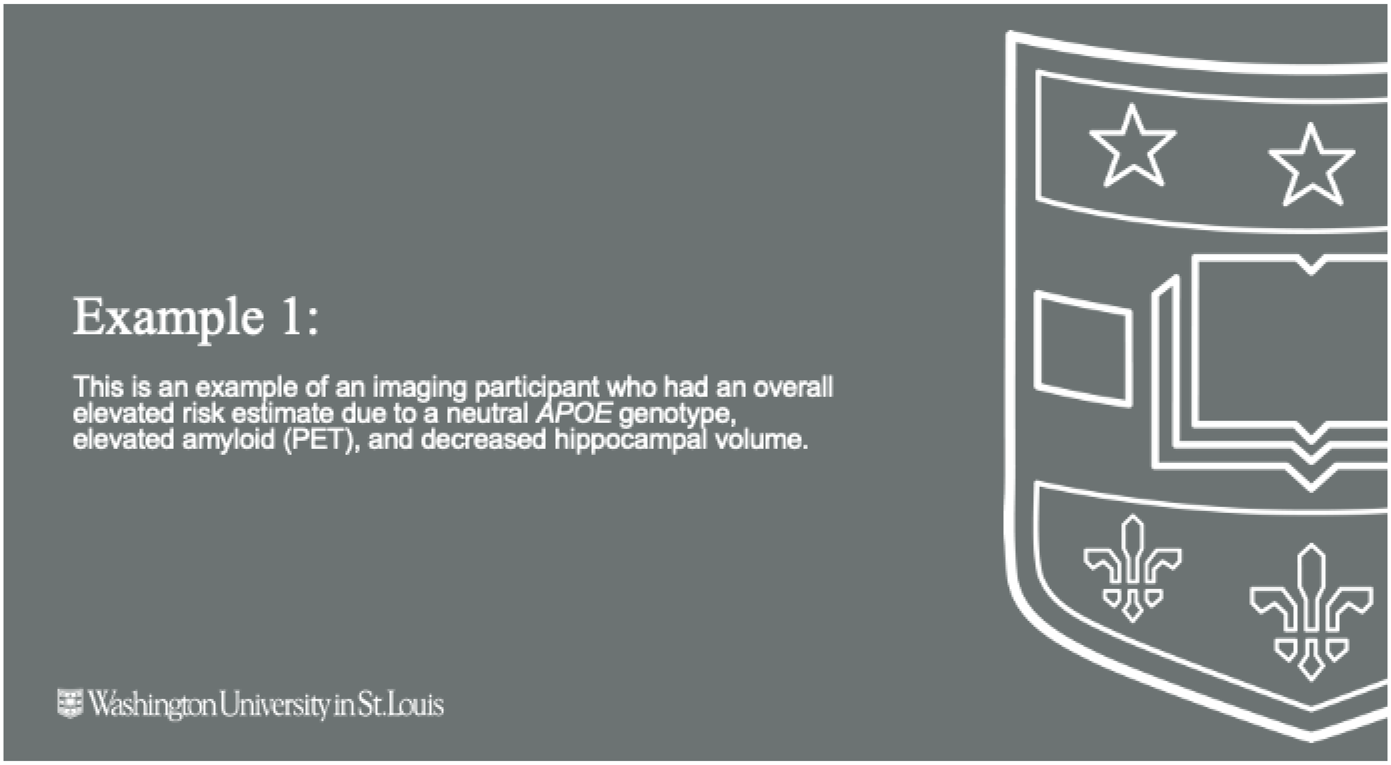
Example 1:
This is an example of an imaging participant who had an overall elevated risk estimate due to a neutral *APOE* genotype, elevated amyloid (PET), and decreased hippocampal volume.
Washington University in St. Louis

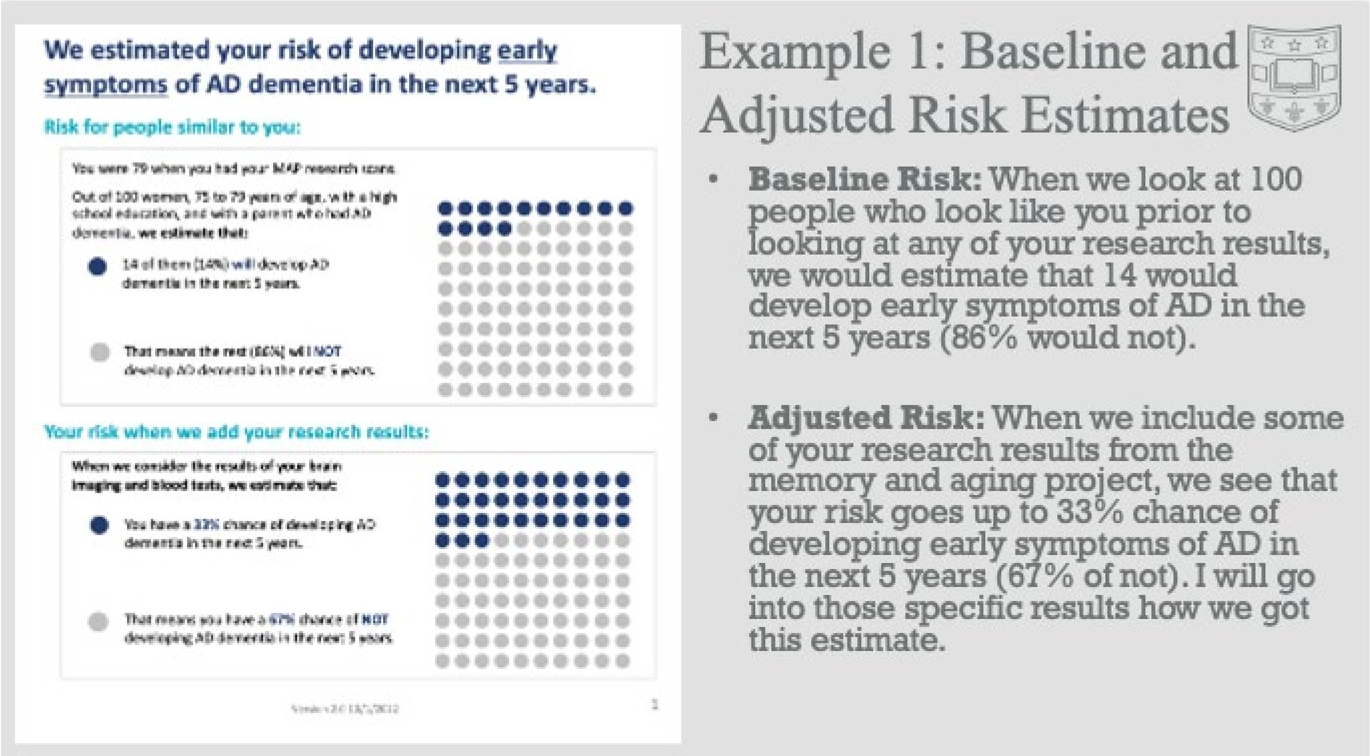
We estimated your risk of developing early symptoms of AD dementia in the next 5 years.
Risk for people similar to you:
14 of them (14%) will develop AD dementia in the next 5 years.
That means the rest (86%) will NOT develop AD dementia in the next 5 years.
Your risk when we add your research results:
When we consider the results of your brain imaging and blood tests, we estimate that:
You have a 33% chance of developing AD dementia in the next 5 years.
That means you have a 67% chance of NOT developing AD dementia in the next 5 years.
Example 1: Baseline and Adjusted Risk Estimates
• **Baseline Risk:** When we look at 100 people who look like you prior to looking at any of your research results, we would estimate that 14 would develop early symptoms of AD in the next 5 years (86% would not).
• **Adjusted Risk:** When we include some of your research results from the memory and aging project, we see that your risk goes up to 33% chance of developing early symptoms of AD in the next 5 years (67% of not). I will go into those specific results how we got this estimate.

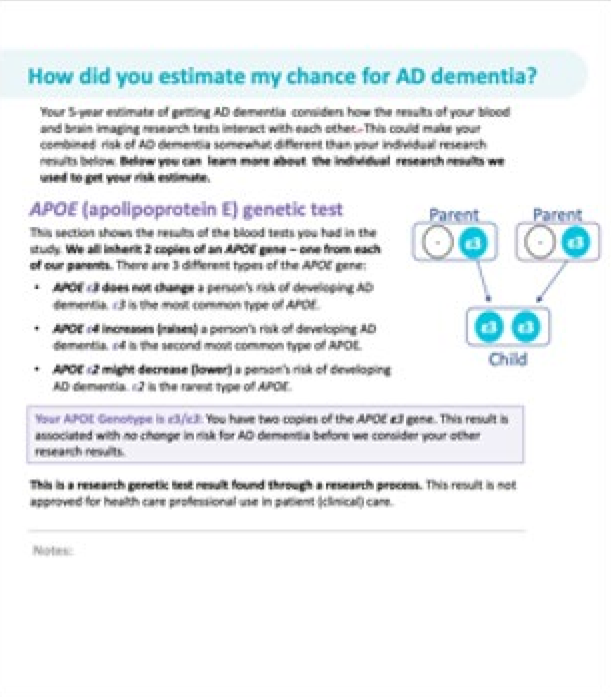

#### How did you estimate my chance for AD dementia?

Your 5-year estimate of getting AD dementia considers how the results of your blood and brain imaging research tests interact with each other. This could make your combined risk of AD dementia somewhat different than your individual research results below. **Below you can learn more about the individual research results we used to get your risk estimate.**

##### *APOE* (apolipoprotein E) genetic test

This section shows the results of the blood tests you had in the study. **We all inherit 2 copies of an *APOE* gene – one from each of our parents.** There are 3 different types of the *APOE* gene:

- ***APOE* ε3 does not change** a person's risk of developing AD dementia. ε3 is the most common type of *APOE*.
- ***APOE* ε4 increases (raises)** a person's risk of developing AD dementia. ε4 is the second most common type of APOE.
- ***APOE* ε2 might decrease (lower)** a person's risk of developing AD dementia. ε2 is the rarest type of *APOE*.

Your APOE Genotype is ε3/ε3: You have two copies of the *APOE ε3* gene. This result is associated with *no change* in risk for AD dementia before we consider your other research results.

**This is a research genetic test result found through a research process.** This result is not approved for health care professional use in patient (clinical) care.

Notes:

### Example 1: *APOE* Genotype

- You may remember having your blood drawn as part of the memory and aging project study. One of the tests that they looked at is a genetic test, looking at a gene called *APOE*.
- Everyone has two copies of the *APOE* gene, one they they inherit from their biological mother and one that they inherit from their biological father.
  - The most common is the ε3 type, which is neutral (does not increase or decrease) risk for developing AD.
  - The second most common is the ε4 type which is known to increase risk for developing AD.
  - The least common is the ε2 type which is known to be protective or decrease risk for developing AD.
- [Point to figure]: In this figure you are the child. You inherited two copies of the common ε3 type, which is associated which does not change your risk of developing AD.
- Any questions?

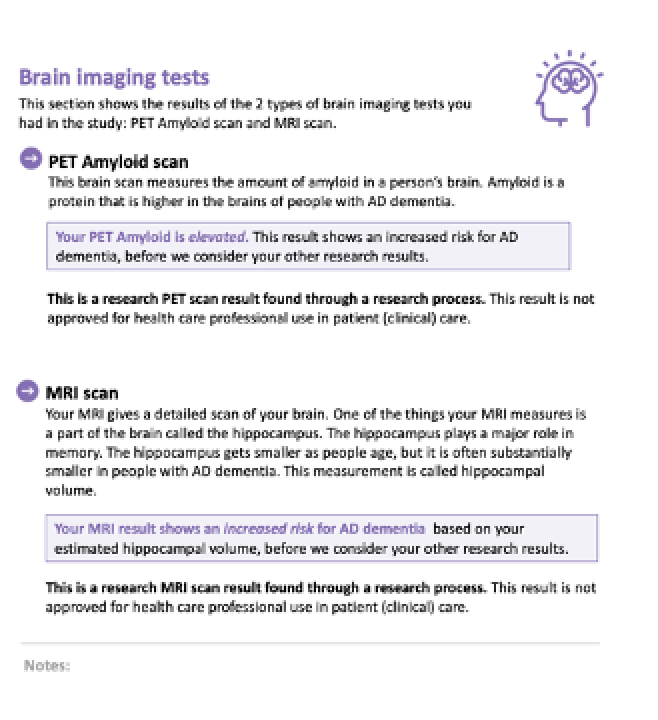

##### Brain imaging tests

This section shows the results of the 2 types of brain imaging tests you had in the study: PET Amyloid scan and MRI scan.

**PET Amyloid scan**

This brain scan measures the amount of amyloid in a person's brain. Amyloid is a protein that is higher in the brains of people with AD dementia.

Your PET Amyloid is *elevated*. This result shows an increased risk for AD dementia, before we consider your other research results.

**This is a research PET scan result found through a research process.** This result is not approved for health care professional use in patient (clinical) care.

**MRI scan**

Your MRI gives a detailed scan of your brain. One of the things your MRI measures is a part of the brain called the hippocampus. The hippocampus plays a major role in memory. The hippocampus gets smaller as people age, but it is often substantially smaller in people with AD dementia. This measurement is called hippocampal volume.

Your MRI result shows an *increased risk* for AD dementia based on your estimated hippocampal volume, before we consider your other research results.

**This is a research MRI scan result found through a research process.** This result is not approved for health care professional use in patient (clinical) care.

Notes:

### Example 1: Amyloid (PET)

- You may remember having a PET scan as part of the memory and aging project.
- One of the scans they did was looking at a protein in the brain called amyloid. Amyloid can build up in the brain as people age, but we know that there tends to be greater levels of amyloid in the brains of people with AD dementia.
- When looking at your PET scan data, we saw that your amyloid levels were elevated, indicating an increased risk for developing AD.
- Elevated amyloid does not guarantee that you will develop AD, but it is associated with increased risk before we consider any of your other research results.
- Questions?

#### Brain imaging tests

This section shows the results of the 2 types of brain imaging tests you had in the study: PET Amyloid scan and MRI scan.

##### PET Amyloid scan

This brain scan measures the amount of amyloid in a person's brain. Amyloid is a protein that is higher in the brains of people with AD dementia.

> Your PET Amyloid is *elevated*. This result shows an increased risk for AD dementia, before we consider your other research results.

**This is a research PET scan result found through a research process.** This result is not approved for health care professional use in patient (clinical) care.

##### MRI scan

Your MRI gives a detailed scan of your brain. One of the things your MRI measures is a part of the brain called the hippocampus. The hippocampus plays a major role in memory. The hippocampus gets smaller as people age, but it is often substantially smaller in people with AD dementia. This measurement is called hippocampal volume.

> Your MRI result shows an *increased risk* for AD dementia based on your estimated hippocampal volume, before we consider your other research results.

**This is a research MRI scan result found through a research process.** This result is not approved for health care professional use in patient (clinical) care.

Notes:

### Example 1: MRI

- You may remember having an MRI scan as part of the memory and aging project.
- One of the things that they look at on the MRI images is called hippocampal volume (HV).
- The hippocampus plays an important role in memory. We know that hippocampal volume size decreases as people age, but tends to be smaller in individuals with AD.
- When looking at your MRI scans, we saw that your HV was decreased, indicating an increased risk for developing AD.
- This means that your HV was smaller than a certain cutoff, NOT that your HV has necessarily decreased from prior scans.
- Decreased HV does not guarantee that you will develop AD, but it is associated with increased risk before we consider any of your other research results.
- Questions?

#### We estimated your risk of developing <u>early symptoms</u> of AD dementia in the next 5 years.

**Risk for people similar to you:**

You were 79 when you had your MAP research scans.

Out of 100 women, 75 to 79 years of age, with a high school education, and with a parent who had AD dementia, **we estimate that:**

- 14 of them (14%) will develop AD dementia in the next 5 years.
- That means the rest (86%) will NOT develop AD dementia in the next 5 years.

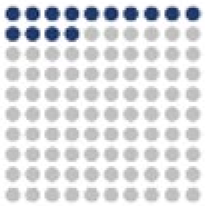

**Your risk when we add your research results:**

**When we consider the results of your brain imaging and blood tests, we estimate that:**

- You have a 33% chance of developing AD dementia in the next 5 years.
- That means you have a 67% chance of NOT developing AD dementia in the next 5 years.

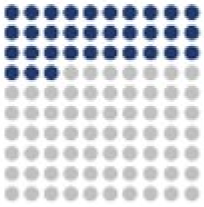

[illegible]

### Example 1: Summary

- When considering these three research results together (neutral *APOE*, increased amyloid, and decreased HV, that is why we see that your risk for AD goes up from 14% to 33%.

##### What should I know about research results?

Make sure to talk to the study team before you share your results with doctors or if you want more information on clinical confirmation. Here's why:

There are different types of health testing. Your doctor orders a **clinical test** to help make healthcare decisions. Clinical tests are certified by the Food and Drug Administration (FDA). A **research test** may not have been certified by the FDA. Research tests may be done in different labs or using different machinery or methods. Research test results are not usually used to make health care decisions. Research tests are not usually placed in your medical record.

We are only returning research results that have scientific evidence about their relationship to AD dementia risk . Before you make any healthcare decisions, it's advised to confirm research test results with an FDA approved clinical test. There may be a cost and insurance implications if the test is clinically confirmed and put in your medical record.

##### What if I have questions about these results later?

If you have questions about these research results after your appointment today, contact the study team:

Amy Oliver, MSW
314-362-6535

You can find more information and resources on AD dementia in the AD dementia brochure you received.

#### Example 1: Disclosure

- I do like to mention that these are research results and not clinical test results. While we are confident in the data we are sharing with you, these tests are not approved by the FDA, and therefore should not be used for making clinical healthcare decisions.
- If you have questions or concerns about your memory or cognition, reach out to your primary care physician so they can discuss clinical testing options with you.
- We will be reaching out to you in a week to check in and see how you are doing with these research results, but if you have any questions before, do not hesitate to reach out.

#### Example 2:

This is an example of an imaging participant who had an overall decreased risk estimate due to a neutral *APOE* genotype, elevated amyloid (PET), and normal hippocampal volume.

Washington University in St. Louis

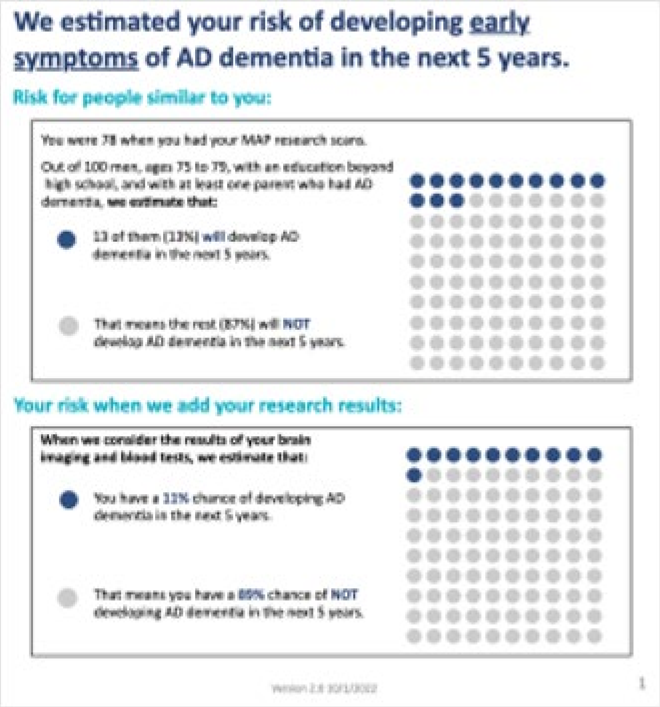

**We estimated your risk of developing early symptoms of AD dementia in the next 5 years.**

**Risk for people similar to you:**

You were 78 when you had your MAP research scans.

Out of 100 men, ages 75 to 79, with an education beyond high school, and with at least one parent who had AD dementia, **we estimate that:**

- 13 of them (13%) will develop AD dementia in the next 5 years.
- That means the rest (87%) will NOT develop AD dementia in the next 5 years.

**Your risk when we add your research results:**

**When we consider the results of your brain imaging and blood tests, we estimate that:**

- You have a 11% chance of developing AD dementia in the next 5 years.
- That means you have a 89% chance of NOT developing AD dementia in the next 5 years.

Version 2.8 10/1/2022 1

### Example 2: Baseline and Adjusted Risk

- **Baseline Risk:** When we look at 100 people who look like you prior to looking at any of your research results, we would estimate that 13 would develop early symptoms of AD in the next 5 years (87% would not).
- **Adjusted Risk:** When we include some of your research results from the memory and aging project, we see that your risk goes down to an 11% chance of developing early symptoms of AD in the next 5 years (89% of not). I will go into those specific results how we got this estimate.

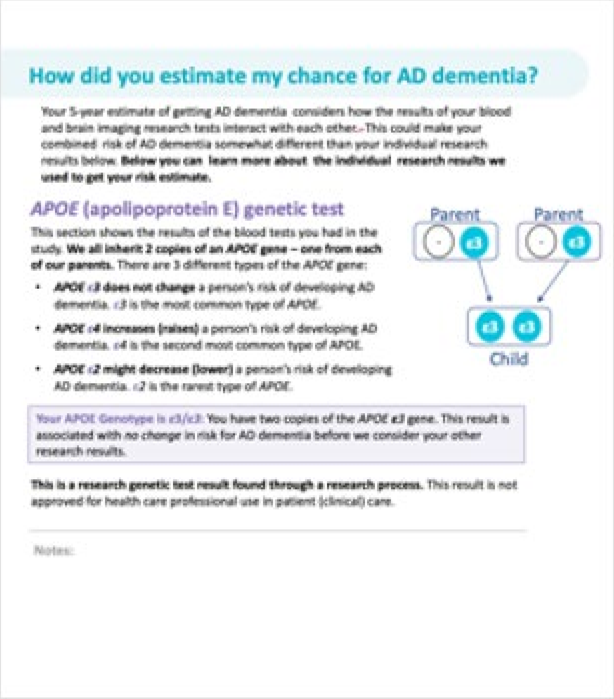

**How did you estimate my chance for AD dementia?**

Your 5-year estimate of getting AD dementia considers how the results of your blood and brain imaging research tests interact with each other. This could make your combined risk of AD dementia somewhat different than your individual research results below. **Below you can learn more about the individual research results we used to get your risk estimate.**

***APOE* (apolipoprotein E) genetic test**

This section shows the results of the blood tests you had in the study. **We all inherit 2 copies of an *APOE* gene – one from each of our parents.** There are 3 different types of the *APOE* gene:

- ***APOE* ε3 does not change** a person's risk of developing AD dementia. ε3 is the most common type of *APOE*.
- ***APOE* ε4 increases (raises)** a person's risk of developing AD dementia. ε4 is the second most common type of APOE.
- ***APOE* ε2 might decrease (lower)** a person's risk of developing AD dementia. ε2 is the rarest type of *APOE*.

Your APOE Genotype is ε3/ε3: You have two copies of the *APOE ε3* gene. This result is associated with *no change* in risk for AD dementia before we consider your other research results.

**This is a research genetic test result found through a research process.** This result is not approved for health care professional use in patient (clinical) care.

Notes:

### Example 2: *APOE* Genotype

- You may remember having your blood drawn as part of the memory and aging project study. One of the tests that they looked at is a genetic test, looking at a gene called *APOE*.
- Everyone has two copies of the *APOE* gene, one they they inherit from their biological mother and one that they inherit from their biological father.
  - The most common is the ε3 type, which is neutral (does not increase or decrease) risk for developing AD.
  - The second most common is the ε4 type which is known to increase risk for developing AD.
  - The least common is the ε2 type which is known to be protective or decrease risk for developing AD.
- [Point to figure]: In this figure you are the child. You inherited two copies of the common ε3 type, which is associated which does not change your risk of developing AD.
- Any questions?

##### Brain imaging tests

This section shows the results of the 2 types of brain imaging tests you had in the study: PET Amyloid scan and MRI scan.

**PET Amyloid scan**

This brain scan measures the amount of amyloid in a person's brain. Amyloid is a protein that is higher in the brains of people with AD dementia.

> Your PET Amyloid is *elevated*. This result shows an increased risk for AD dementia, before we consider your other research results.

**This is a research PET scan result found through a research process.** This result is not approved for health care professional use in patient (clinical) care.

**MRI scan**

Your MRI gives a detailed scan of your brain. One of the things your MRI measures is a part of the brain called the hippocampus. The hippocampus plays a major role in memory. The hippocampus gets smaller as people age, but it is often substantially smaller in people with AD dementia. This measurement is called hippocampal volume.

> Your MRI result shows a *decreased risk* for AD dementia based on your estimated hippocampal volume, before we consider your other research results.

**This is a research MRI scan result found through a research process.** This result is not approved for health care professional use in patient (clinical) care.

Notes:

#### Example 2: Amyloid (PET)

- You may remember having a PET scan as part of the memory and aging project.
- One of the scans they did was looking at a protein in the brain called amyloid. Amyloid can build up in the brain as people age, but we know that there tends to be greater levels of amyloid in the brains of people with AD dementia.
- When looking at your PET scan data, we saw that your amyloid levels were elevated, indicating an increased risk for developing AD.
- Elevated amyloid does not guarantee that you will develop AD, but it is associated with increased risk before we consider any of your other research results.
- Questions?

##### Brain imaging tests

This section shows the results of the 2 types of brain imaging tests you had in the study: PET Amyloid scan and MRI scan.

**PET Amyloid scan**

This brain scan measures the amount of amyloid in a person's brain. Amyloid is a protein that is higher in the brains of people with AD dementia.

> Your PET Amyloid is *elevated*. This result shows an increased risk for AD dementia, before we consider your other research results.

**This is a research PET scan result found through a research process.** This result is not approved for health care professional use in patient (clinical) care.

**MRI scan**

Your MRI gives a detailed scan of your brain. One of the things your MRI measures is a part of the brain called the hippocampus. The hippocampus plays a major role in memory. The hippocampus gets smaller as people age, but it is often substantially smaller in people with AD dementia. This measurement is called hippocampal volume.

> Your MRI result shows a *decreased risk* for AD dementia based on your estimated hippocampal volume, before we consider your other research results.

**This is a research MRI scan result found through a research process.** This result is not approved for health care professional use in patient (clinical) care.

Notes:

#### Example 2: MRI

- You may remember having an MRI scan as part of the memory and aging project.
- One of the things that they look at on the MRI images is called hippocampal volume (HV).
- The hippocampus plays an important role in memory. We know that hippocampal volume size decreases as people age, but tends to be smaller in individuals with AD.
- When looking at your MRI scans, we saw that your HV was NOT decreased, indicating an decreased risk for developing AD before looking at any of your other research results.
- Questions?

**We estimated your risk of developing <u>early symptoms</u> of AD dementia in the next 5 years.**

**Risk for people similar to you:**

You were 78 when you had your MAP research scans.

Out of 100 men, ages 75 to 79, with an education beyond high school, and with at least one parent who had AD dementia, **we estimate that:**

- 13 of them (13%) **will** develop AD dementia in the next 5 years.
- That means the rest (87%) will **NOT** develop AD dementia in the next 5 years.

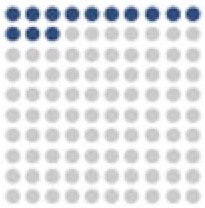

**Your risk when we add your research results:**

**When we consider the results of your brain imaging and blood tests, we estimate that:**

- You have a 11% chance of developing AD dementia in the next 5 years.
- That means you have a 89% chance of **NOT** developing AD dementia in the next 5 years.

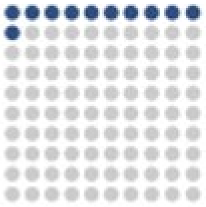

Version 2.8 10/1/2022 1

### Example 2: Summary

- When we look at all of your research results together, despite having elevated amyloid, because of the neutral *APOE*, and normal HV, your risk is actually decreased from 13% to 11%.
- Questions?

#### What should I know about research results?

Make sure to talk to the study team before you share your results with doctors or if you want more information on clinical confirmation. Here's why:

There are different types of health testing. Your doctor orders a **clinical test** to help make healthcare decisions. Clinical tests are certified by the Food and Drug Administration (FDA). A **research test** may not have been certified by the FDA. Research tests may be done in different labs or using different machinery or methods. Research test results are not usually used to make health care decisions. Research tests are not usually placed in your medical record.

We are only returning research results that have scientific evidence about their relationship to AD dementia risk . Before you make any healthcare decisions, it's advised to confirm research test results with an FDA approved clinical test. There may be a cost and insurance implications if the test is clinically confirmed and put in your medical record.

#### What if I have questions about these results later?

If you have questions about these research results after your appointment today, contact the study team:

Amy Oliver, MSW
314-362-6535

You can find more information and resources on AD dementia in the AD dementia brochure you received.

### Example 2: Disclosure

- I do like to mention that these are research results and not clinical test results. While we are confident in the data we are sharing with you, these tests are not approved by the FDA, and therefore should not be used for making clinical healthcare decisions.
- If you have questions or concerns about your memory or cognition, reach out to your primary care physician so they can discuss clinical testing options with you.
- We will be reaching out to you in a week to check in and see how you are doing with these research results, but if you have any questions before, do not hesitate to reach out.

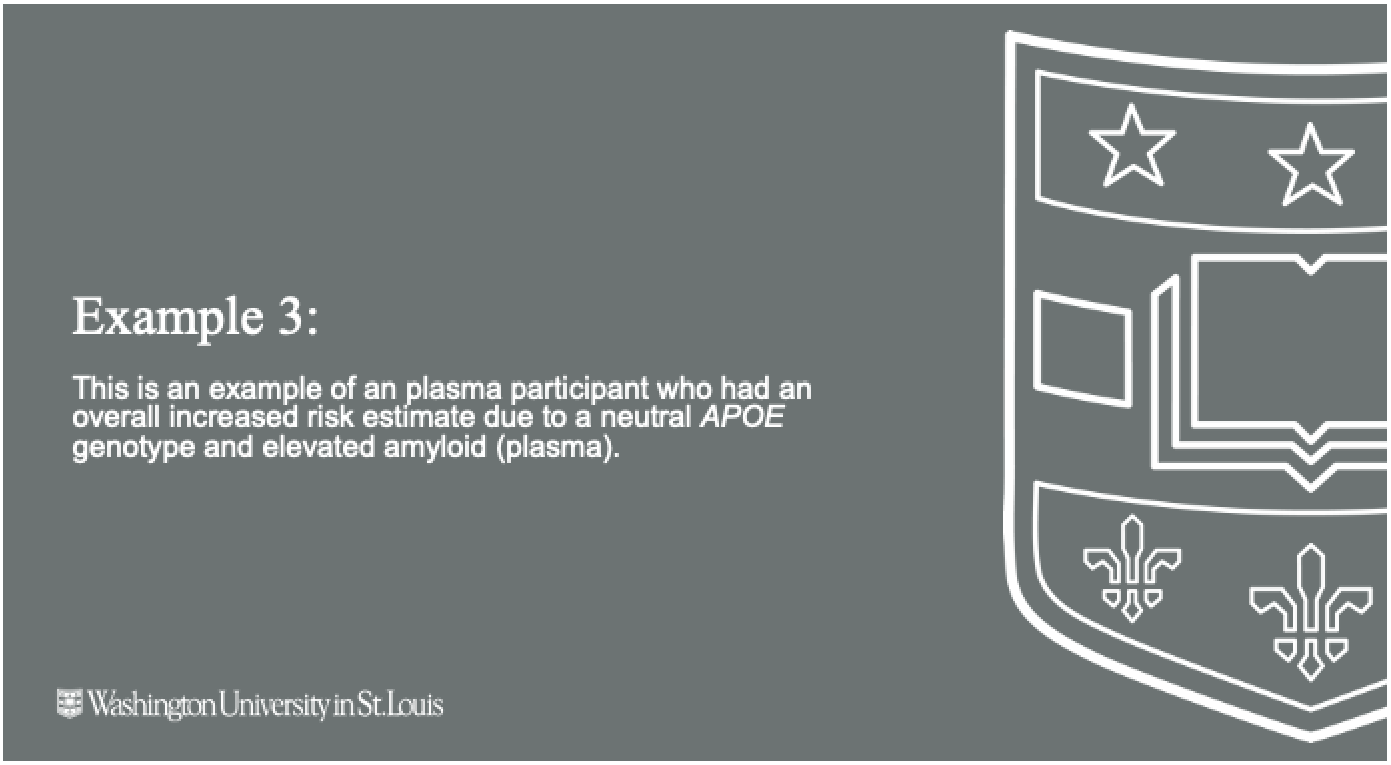
Example 3:
This is an example of an plasma participant who had an overall increased risk estimate due to a neutral APOE genotype and elevated amyloid (plasma).
Washington University in St. Louis

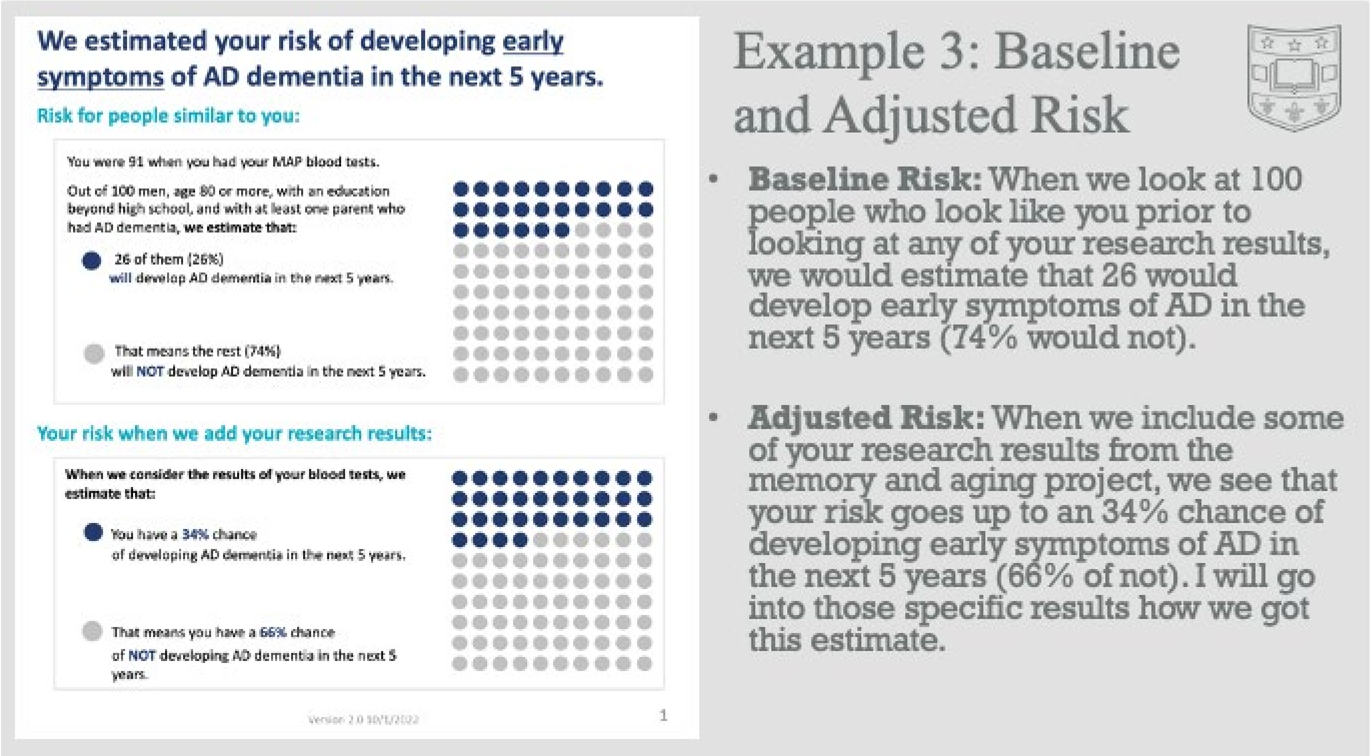
We estimated your risk of developing early symptoms of AD dementia in the next 5 years.
Risk for people similar to you:
You were 91 when you had your MAP blood tests.
Out of 100 men, age 80 or more, with an education beyond high school, and with at least one parent who had AD dementia, we estimate that:
26 of them (26%) will develop AD dementia in the next 5 years.
That means the rest (74%) will NOT develop AD dementia in the next 5 years.
Your risk when we add your research results:
When we consider the results of your blood tests, we estimate that:
You have a 34% chance of developing AD dementia in the next 5 years.
That means you have a 66% chance of NOT developing AD dementia in the next 5 years.
Version 2.0 10/1/2022
1
Example 3: Baseline and Adjusted Risk
Baseline Risk: When we look at 100 people who look like you prior to looking at any of your research results, we would estimate that 26 would develop early symptoms of AD in the next 5 years (74% would not).
Adjusted Risk: When we include some of your research results from the memory and aging project, we see that your risk goes up to an 34% chance of developing early symptoms of AD in the next 5 years (66% of not). I will go into those specific results how we got this estimate.

#### How did you estimate my chance for AD dementia?

Your 5-year estimate of getting AD dementia considers how the results of your two blood tests, *APOE* genetic test and plasma amyloid test, interact with each other. This could make your combined risk of AD dementia somewhat different than your individual research results below. **Below you can learn more about the individual research results we used to get your risk estimate.**

##### *APOE* (apolipoprotein E) Genetic Test

This section shows the results of the blood tests you had in the study. **We all inherit 2 copies of an *APOE* gene – one from each of our parents.** There are 3 different types of the *APOE* gene:

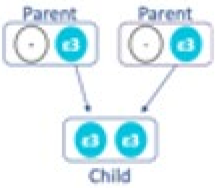

- ***APOE* ε3 does not change** a person's risk of developing AD dementia. ε3 is the most common type of *APOE*.
- ***APOE* ε4 increases (raises)** a person's risk of developing AD dementia. ε4 is the second most common type of APOE.
- ***APOE* ε2 might decrease (lower)** a person's risk of developing AD dementia. ε2 is the rarest type of *APOE*.

Your *APOE* genotype is ε3/ε3. You have one copy of the *APOE* ε3 gene and one copy of the *APOE* ε3 gene. This result is associated with *no change* in risk for AD dementia before we consider your other research results.

**This is a research genetic test result found through a research process.** This result is not approved for health care professional use in patient (clinical) care.

Notes:

### Example 3: *APOE* Genotype

- You may remember having your blood drawn as part of the memory and aging project study. One of the tests that they looked at is a genetic test, looking at a gene called *APOE*.
- Everyone has two copies of the *APOE* gene, one they they inherit from their biological mother and one that they inherit from their biological father.
  - The most common is the ε3 type, which is neutral (does not increase or decrease) risk for developing AD.
  - The second most common is the ε4 type which is known to increase risk for developing AD.
  - The least common is the ε2 type which is known to be protective or decrease risk for developing AD.
- [Point to figure]: In this figure you are the child. You inherited two copies of the common ε3 type, which is associated which does not change your risk of developing AD.
- Any questions?

#### Amyloid Blood Test

This section shows the results of the blood test you had for this study.

**Amyloid Blood Test**

Amyloid, a protein that builds up in the brains of people with AD dementia, is also found in small amounts in the blood. The amount and types of amyloid found in the blood can tell us if amyloid is building up in the brain.

Your amyloid blood test shows an *increased risk* for AD dementia, before we consider your other research results.

**This is a research amyloid blood test result found through a research process.** This result is not approved for health care professional use in patient (clinical) care.

### Example 3: Amyloid (Plasma)

- One of the other things that they were looking at in your blood a protein in the brain called amyloid. Amyloid can build up in the brain as people age, but we know that there tends to be greater levels of amyloid in the brains of people with AD dementia.
- When looking at your data, we saw that your amyloid levels were elevated, indicating an increased risk for developing AD
- Elevated amyloid does not guarantee that you will develop AD, but it is associated with increased risk before we consider any of your other research results.
- Questions?

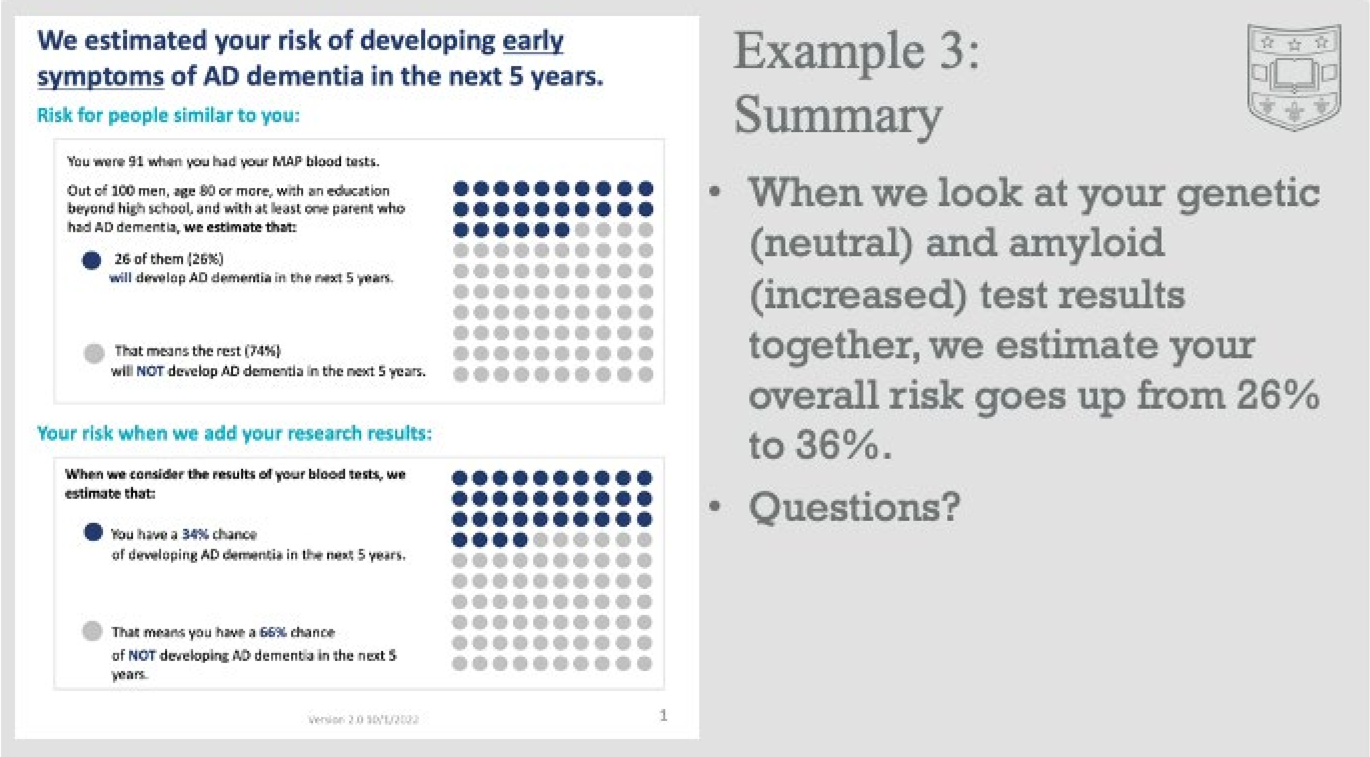
We estimated your risk of developing early symptoms of AD dementia in the next 5 years.
Risk for people similar to you:
You were 91 when you had your MAP blood tests.
Out of 100 men, age 80 or more, with an education beyond high school, and with at least one parent who had AD dementia, we estimate that:
26 of them (26%) will develop AD dementia in the next 5 years.
That means the rest (74%) will NOT develop AD dementia in the next 5 years.
Your risk when we add your research results:
When we consider the results of your blood tests, we estimate that:
You have a 34% chance of developing AD dementia in the next 5 years.
That means you have a 66% chance of NOT developing AD dementia in the next 5 years.
1
Example 3: Summary
When we look at your genetic (neutral) and amyloid (increased) test results together, we estimate your overall risk goes up from 26% to 36%.
Questions?

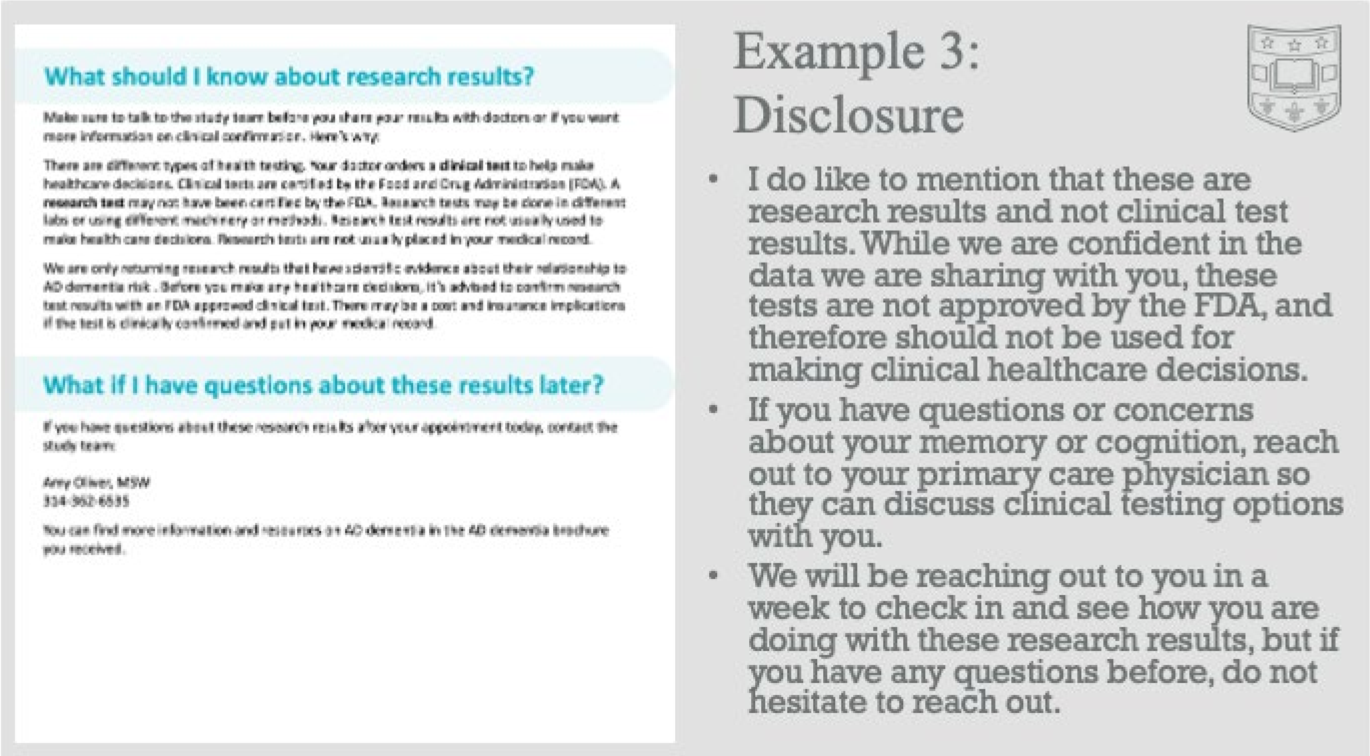
What should I know about research results?
Make sure to talk to the study team before you share your results with doctors or if you want more information on clinical confirmation. Here's why:
There are different types of health testing. Your doctor orders a clinical test to help make healthcare decisions. Clinical tests are certified by the Food and Drug Administration (FDA). A research test may not have been certified by the FDA. Research tests may be done in different labs or using different machinery or methods. Research test results are not usually used to make health care decisions. Research tests are not usually placed in your medical record.
We are only returning research results that have scientific evidence about their relationship to AD dementia risk. Before you make any healthcare decisions, it's advised to confirm research test results with an FDA approved clinical test. There may be a cost and insurance implications if the test is clinically confirmed and put in your medical record.
What if I have questions about these results later?
If you have questions about these research results after your appointment today, contact the study team:
Amy Oliver, MSW
You can find more information and resources on AD dementia in the AD dementia brochure you received.
Example 3: Disclosure
I do like to mention that these are research results and not clinical test results. While we are confident in the data we are sharing with you, these tests are not approved by the FDA, and therefore should not be used for making clinical healthcare decisions.
If you have questions or concerns about your memory or cognition, reach out to your primary care physician so they can discuss clinical testing options with you.
We will be reaching out to you in a week to check in and see how you are doing with these research results, but if you have any questions before, do not hesitate to reach out.

#### Wrapping up the session

Washington University in St. Louis

#### Things you may want to ask…

- How do you feel about these results?
- Have you thought about who/if you may want to share these results with anyone?
- How would you explain these results to a family member/friend?
- What other questions do you have?

#### Teach Back Method

- The teach-back method is an evidence-based way to assess patient understanding.
- This process can sometimes be awkward and feel like you are quizzing the participant.
- Here is some language that can be used for effective teach-back:
  - "How would you explain these results with a family member or friend?"
  - "I want to make sure that I explained things well. Can you explain what we discussed in your own words, so I can make sure that I did?"
  - We have talked about a lot of things just now—can you tell me in your own words what we discussed?"
  - "What other questions do you have?"

#### FAQs

Washington University in St. Louis

### FAQ

- **Are there any drugs available for asymptomatic individuals with elevated amyloid?**
  - Currently no drugs FDA approved for asymptomatic patients
  - Ask your PCP about it if it is something you are interested in incase there are new drugs that become available in the future.
- **Are there any clinical trials for asymptomatic individuals with elevated amyloid?**
  - AHEAD Study: Double blind placebo control randomized clinical trial of lecanemab (trade name Leqembi) drug that was recently FDA approved for mild AD
  - Eligibility Requirements:
    - CN (CDR of 0), Elevated Amyloid
  - Recruitment content info: Sonia Simons
- **Are these research results ever put in my health records?**
  - We will not share your research results with your doctor(s). If you choose to share this information with your doctor(s), they may place it in your medical record. This may negatively affect future purchases of life or long-term care insurance. Make sure to discuss the implications before sharing this information with your doctor(s).
  - The Genetic Information Nondiscrimination Act (GINA) is a federal law that protects against genetic discrimination in health insurance and employment. GINA does not protect against discrimination in disability, life, or long-term care insurance. There are no laws currently that protect against discrimination based on brain imaging results.
- **Is there a way to know which *APOE* allele came from which parent?**
  - Not without testing the parents

### FAQ

- **Is there a way to predict how amyloid will change?**
  - Not from this data
- **Is there a reason why HV would be decreased other than AD?**
  - Age
  - We are not using longitudinal data, so may not be a change, but rather just smaller than average
- **Are these results AD specific or apply to other forms of dementia (ie: FTD or LBD)?**
  - *APOE*: AD specific
  - Amyloid: AD specific. Other forms of dementia are associated with different types of pathology.
  - Hippocampal Volume: Unclear
- **How much more accurate is a PET scan compared to the PrecivityAD2 blood test at measuring amyloid on the brain?**
  - PET Amyloid test is better, but the blood test is still good. They both measure the amount of amyloid on the brain. The blood test uses newer technology that is not quite as accurate as PET. This is why when imaging if available, we use that. The new blood test allows us to return results to participants who do not/cannot have imaging.
